# Whole genome sequencing delineates regulatory and novel genic variants in childhood cardiomyopathy

**DOI:** 10.1101/2020.10.12.20211474

**Authors:** Robert Lesurf, Abdelrahman Said, Oyediran Akinrinade, Jeroen Breckpot, Kathleen Delfosse, Ting Liu, Roderick Yao, Fintan McKenna, Ramil R. Noche, Winona Oliveros, Kaia Mattioli, Anastasia Miron, Qian Yang, Guoliang Meng, Michelle Chan Seng Yue, Wilson WL Sung, Bhooma Thiruvahindrapuram, Genomics England Research Consortium, Jane Lougheed, Erwin Oechslin, Lynn Bergin, John Smythe, Tapas Mondal, Marta Melé, Philipp G. Maass, James Ellis, Stephen W. Scherer, Seema Mital

## Abstract

Cardiomyopathy (CMP) is a heritable genetic disorder. Protein-coding variants account for 20-30% of cases. The contribution of variants in non-coding DNA elements that regulate gene expression has not been explored. We performed whole-genome sequencing (WGS) of 228 unrelated CMP families. Besides pathogenic protein-coding variants in known CMP genes, 5% cases harbored rare loss-of-function variants in novel cardiac genes, with *NRAP* and *FHOD3* being strong candidates. WGS also revealed a high burden of high-risk variants in promoters and enhancers of CMP genes in an additional 20% cases (Odds ratio 2.14, 95% CI 1.60-2.86, p=5.26×10^−7^ vs 1326 controls) with genes involved in α-dystroglycan glycosylation (*FKTN, DTNA*) and desmosomal signaling (*DSC2, DSG2*) specifically enriched for regulatory variants (False discovery rate <0.03). These findings were independently replicated in the Genomics England CMP cohort (n=1266). The functional effect of non-coding variants on transcription was functionally validated in patient myocardium and reporter assays in human cardiomyocytes, and that of novel gene variants in zebrafish knockouts. Our results show that functionally active variants in novel genes and in regulatory elements of CMP genes contribute strongly to the genomic etiology of childhood-onset CMP.

## INTRODUCTION

Cardiomyopathy (CMP) is a genetic disease of heart muscle with a prevalence of 1:500 to 1:2500 in the general population (depending on CMP type). Over 20 million people worldwide are estimated to be living with the disease^1^. The actual prevalence is estimated to be even higher given that many patients who carry a gene defect have not manifested the disease yet. Several thousand new cases of CMP are diagnosed each year in North America alone^2^. Over a third of the cases are inherited while the remainder are sporadic^3^. Majority are autosomal dominant in nature caused by rare damaging variants in genes that impact muscle structure and function^4,5^. There are five phenotypes – hypertrophic (HCM), dilated (DCM), restrictive (RCM), left ventricular non-compaction cardiomyopathy (LVNC) and arrhythmogenic ventricular cardiomyopathy (AVC). The disease has a high penetrance in childhood^6^ with CMP being the leading cause of heart failure and sudden cardiac death in children^7^. The greater severity of disease in childhood onset CMP is presumed to be related in part to genetic differences that have not been systematically evaluated^8^.

There is considerable genetic overlap between different CMP subtypes. While sarcomeric genes including *MYH7* and *MYBPC3*, explain ∼50% of all HCM cases, other CMPs are more polygenic, and despite the inclusion of upwards of 100 putative CMP disease genes in clinical diagnostic testing panels, over 70% of CMPs remain gene-elusive (including familial cases)^9–11^. This is in part because the standard gene panel tests typically only capture small sequence-level variants within the coding regions of known CMP genes, and miss hard to sequence regions, most intronic splicing events, structural variation, and novel genes not included on the panels. Importantly, these tests do not evaluate the non-coding genome that harbors DNA regulatory sequences including core and proximal promoters and enhancers, as well as distal regulatory elements^12^. These variants can disrupt the transcriptional activation process through multiple mechanisms including alterations in chromatin structure, non-coding RNA, transcript stability, and importantly, through the alteration of the DNA sequence of transcription factor binding sites (TFBS).

A growing number of whole genome sequencing (WGS) studies are identifying novel genetic variants in pediatric and familial disease^13–15^. In autism spectrum disorder, a complex genetic condition, WGS has enabled identification of putative non-coding regions as hotspots for *de novo* germline variants^16,17^, new candidate risk genes^18^, and novel mechanisms of mutation only discovered by WGS^19,20^. More recently, WGS identified a higher burden of *de novo* variants in the enhancers of disease-associated genes in congenital heart disease patients compared with controls^21^. However, only 5 of the 31 enhancers identified were associated with altered transcription levels of the target genes. Compared to CHD which is a complex disorder that includes not only genetic but also environmental causes, the role of regulatory variants has not been explored in CMP, a primarily genetic disorder.

Here, we used WGS to characterize all classes of genetic variation in a unique and exhaustively-phenotyped cohort ascertained for childhood-onset CMP. WGS identified 11 novel genes important for clinical diagnostic testing, and also identified a significantly higher burden of regulatory variants in CMP genes in 20% of cases compared to controls, findings that were replicated in an independent CMP cohort. The function of the most important variants identified was confirmed through a study of endogenous gene expression in patient myocardium, human cell-based assays^22,23^ and CRISPR gene editing of zebrafish embryos, providing a paradigm for WGS interpretation in future genomic studies of childhood-onset CMP as well as other genetic disorders.

## RESULTS

### Variant yield on WGS in the discovery cohort

The *discovery cohort* consisted of 228 unrelated probands <21 years old at diagnosis with primary CMP, and 69 affected and unaffected family members (**Supplementary Table 1**). The cohort included 49% DCM, 33% HCM, 7% LVNC, 6% RCM, and 3% AVC. 29% of cases had a positive family history of CMP. WGS was performed on genomic DNA using Illumina HiSeq X platform at a median sequencing coverage of 31X (range: 20-50X). We interrogated 133 genes represented in different commercial CMP gene panels (**Supplementary Table 2**) for rare [population minor allele frequency (MAF) <0.01%], predicted damaging missense, loss-of-function (LoF) (frameshift, stopgain/stoploss, splicing) and high-risk regulatory variants. **Figure 1a** depicts the workflow for filtering pathogenic and likely pathogenic protein-coding single nucleotide variants (SNVs), insertion-deletions (indels), copy number variants (CNVs), and high-risk regulatory variants in the overall cohort. Protein-coding variants were classified as pathogenic (including likely pathogenic) using the American College of Medical Genetics (ACMG) criteria^17,24–30^. Pathogenic protein-coding SNVs and indels in known CMP genes were detected in 78/228 (34%) cases, and CNVs in 6/228 (2%) cases. Only two cases harbored homozygous variants. An additional 20% cases harbored high-risk variants in regulatory elements of CMP genes, and 5% cases harbored likely pathogenic LoF variants in novel candidate genes (**Figure 1b**). Variant distribution by CMP subtype, by patient and by gene category is shown in **Figures 1c-e**.

**Figure 1:**
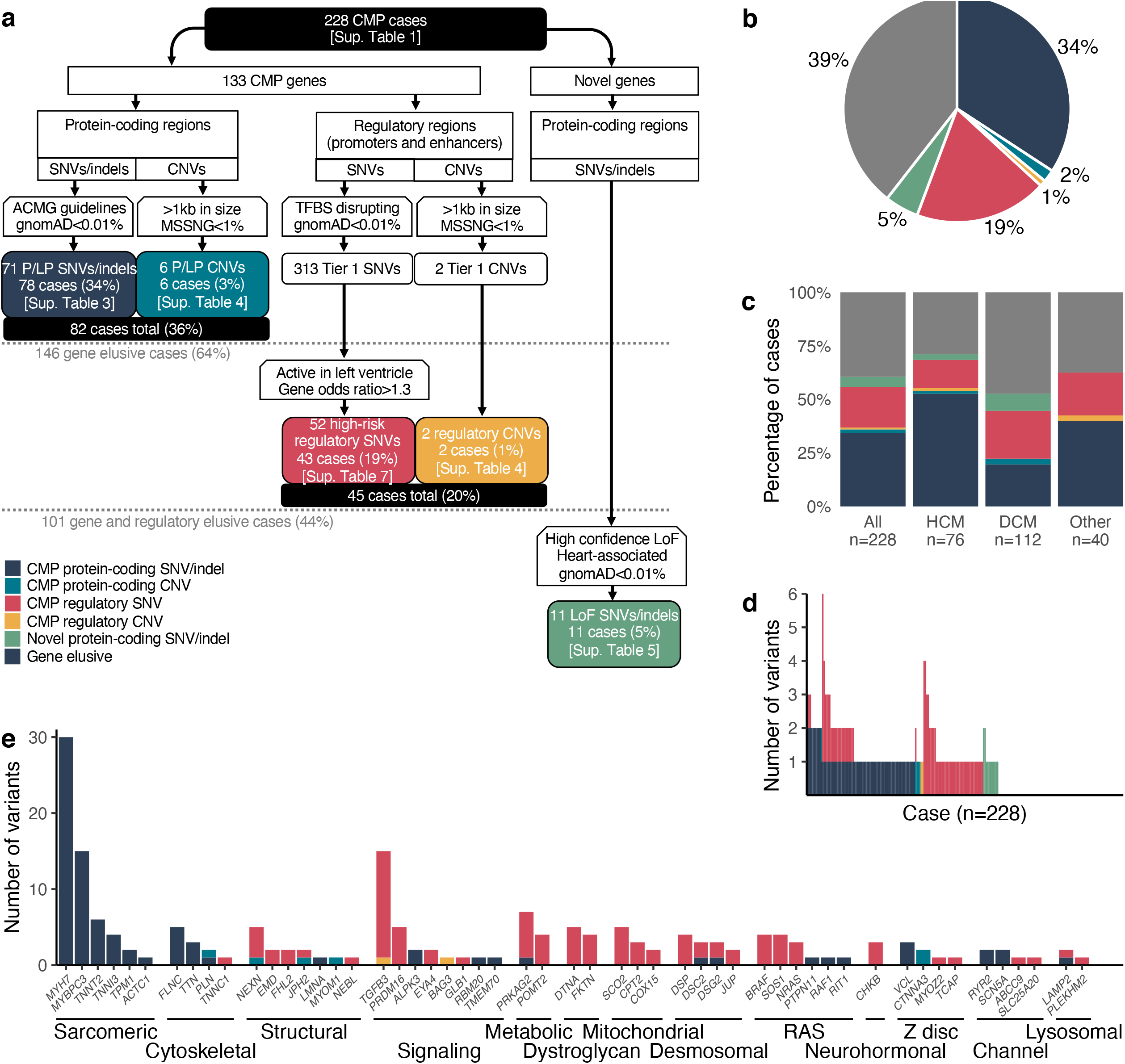
Yield of protein-coding and regulatory variants in 228 unrelated childhood CMP cases. **(a)** Flow-chart showing the selection process and yield of protein-coding and regulatory variants in the overall cohort and in the gene-elusive subset. 36% of all cases harbored at least one pathogenic protein-coding variant in a CMP gene; among the remaining 146 gene elusive cases, 20% harbored at least one high risk regulatory variant in a CMP gene; and an additional 5% harbored a LoF variant in a novel gene. (**b**) Pie diagram showing the distribution of protein-coding and regulatory variants in CMP genes and LoF variants in novel genes across the cohort (n=228). WGS identified putatively causal protein-coding SNVs/indels/CNVs in CMP genes in 36% of cases, high risk variants in regulatory elements of CMP genes in an additional 20% of cases, and loss of function (LoF) variants in novel genes in an additional 5% of cases. (**c**) Variant distribution by CMP subtypes: HCM cases had a higher yield of pathogenic protein-coding variants compared to other CMP subtypes (OR 3.14, CI 1.77-5.57, p=1.22×10^−4^). (**d**) Variant burden by patient in the cohort: 11 cases (5%) had multiple protein-coding variants in known CMP genes, 10 cases (4%) had multiple regulatory variants, and 23 cases (10%) had both protein-coding and regulatory variants in CMP genes. (**e**) Variant distribution by functional gene categories: Of all the pathogenic protein-coding variants, 64% were in sarcomeric genes which represented a significant enrichment compared to other gene categories (binomial p=3.16×10^−49^). Conversely, none of the high-risk regulatory variants were in sarcomeric genes. CMP, cardiomyopathy; SNV, single nucleotide variant; CNV, copy number variant; gnomAD, Genome Aggregation Database; ACMG, American College of Medical Genetics; TFBS, transcription factor binding site; P/LP, pathogenic or likely pathogenic; LoF, loss of function; HCM, hypertrophic cardiomyopathy; DCM, dilated cardiomyopathy;

### Protein-coding variants in known CMP genes

Protein-coding SNVs and CNVs are described in **Supplementary Tables 3 & 4**. The majority (64%) of pathogenic protein-coding variants were in sarcomeric genes which represented a significant enrichment compared to other gene categories (binomial p=3.16×10^−49^). HCM cases had a higher yield of pathogenic protein-coding variants compared to other CMP subtypes with odds ratio (OR) 3.14, 95% confidence intervals (CI) 1.77-5.57 (p=1.22×10^−4^). Of note, WGS detected pathogenic protein-coding variants in 17/228 (7.5%) cases previously missed by panel-based clinical genetic testing since not all CMP genes are captured by commercial testing panels, and none of the gene panels explore for CNVs^31^.

#### Effect of protein-coding variants on myocardial expression of target genes

A unique feature of our biobank is access to myocardial samples from patients undergoing surgery or cardiac transplantation. RNA sequencing was performed in LV myocardial samples from 35 sequenced CMP patients to validate the effect of LoF SNVs and CNVs on gene expression. **Figure 2a-c** shows that candidate gene mRNA expression was below the 25^th^ percentile in the myocardium of patients harbouring LoF SNVs (*DSC2, FLNC, MYBPC3*) compared to the remaining cohort. Endogenous gene expression levels were also reduced in patients with single copy deletion CNVs impacting both the promoter and first exons of the genes *JPH2* and *NEXN*, and exon 11 of *CTNNA3* (**Figure 2d-f**). The ability to show the impact of coding variants on endogenous gene expression in the target organ is a unique finding that supports the use of patient myocardium to validate variant pathogenicity.

**Figure 2:**
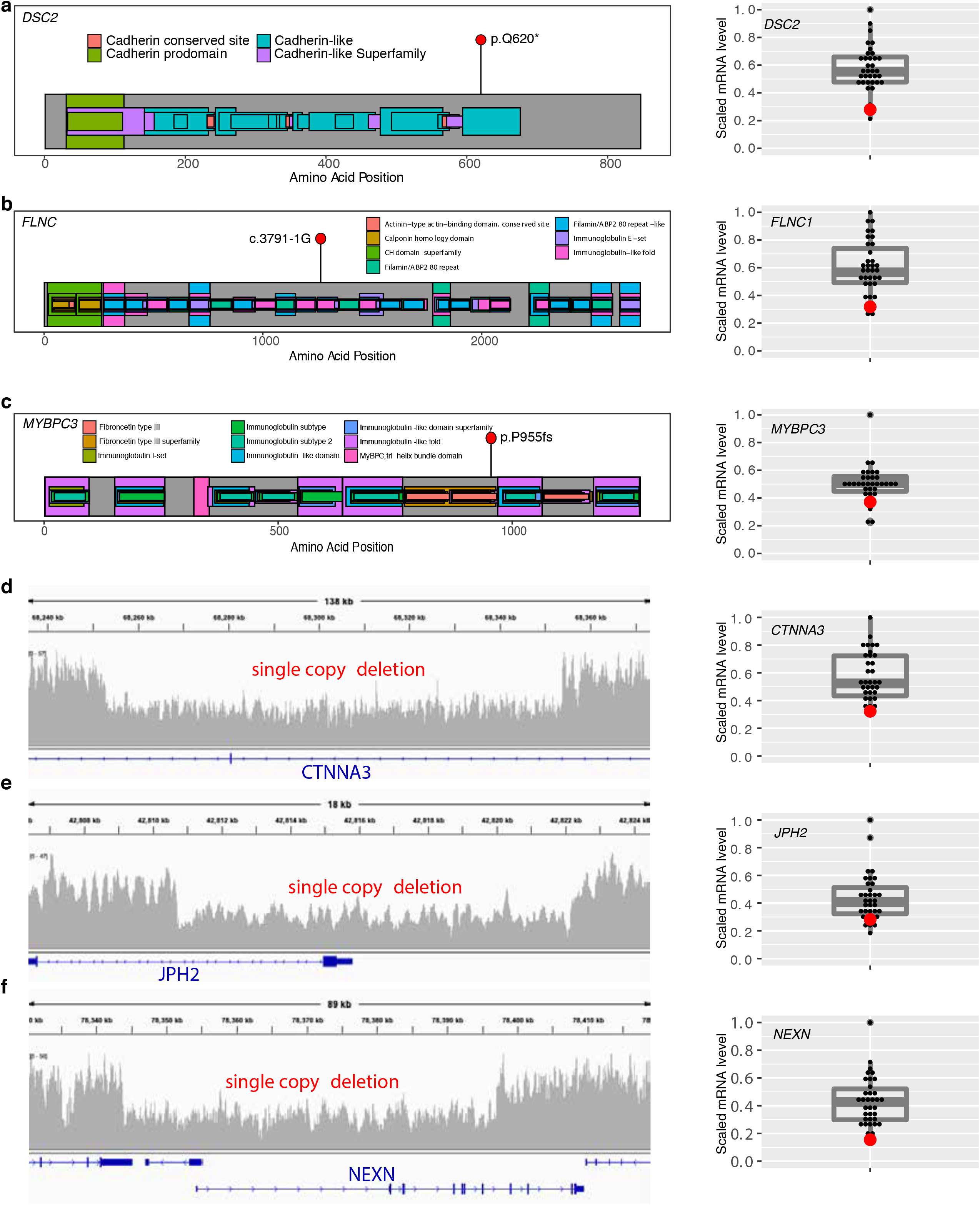
Effect of loss of function and copy number deletions in CMP genes on myocardial gene expression. The figure shows LV myocardial gene expression using RNA sequencing in the patient harboring a loss of function or copy number deletion (red dot) compared to other cases without the variant (grey dots) (n=35 cases). (**a-c**) Left panels show the amino acid position of three pathogenic loss of function variants in *DSC2* (stopgain), *FLNC* (splice acceptor), *MYBPC3* (frameshift deletion) predicted to result in nonsense-mediated decay of mRNA. The right panels show scaled RPKM expression of target mRNA to below the 25^th^ percentile compared to the remaining cohort; (**d-f**) The left panels show the genomic location of three single CNV deletions in *CTNNA3, JPH2, NEXN* genes. The right panels show scaled RPKM expression of target mRNA to below the 25^th^ percentile compared to the remaining cohort. RPKM, Reads Per Kilobase of transcript, per Million mapped reads

### Protein-coding LoF variants in novel CMP genes

WGS provided us an opportunity to explore for novel biologically relevant genes beyond known CMP genes as potential sources of pathogenic coding variants. We searched for rare (gnomAD MAF<0.01%) predicted deleterious heterozygous and homozygous LoF variants in genes involved in heart function with moderate-high heart expression, that were deemed to be constrained for LoF variants^32,33^. Using these criteria, we identified rare LoF variants in 11 novel genes in CMP patients who did not harbor a pathogenic protein-coding variant (5% of the cohort) (**Supplementary Table 5**). Exploration of 1266 independent CMP probands in the 100,000 Genomes Project replication cohort identified rare heterozygous or homozygous LoF variants in five of these novel genes (*FHOD3, NRAP, PDE4DIP, PTGDS*, and *TRPM4*). *FHOD3* harbored the highest proportion of LoF variants, with variants identified in five additional CMP cases from the 100,000 Genomes Project as opposed to only one ICGC control sample.

Of these 11 genes, we explored *FHOD3* and *NRAP* further as strong candidates since they are known to have high heart-specific expression^34,35^, are important in the maintenance of sarcomeric and actin cytoskeleton in the heart, and have been associated with CMP in mouse studies and in small case series^36–43^. In our cohort, we found a rare frameshift variant in *FHOD3* in a DCM patient and a rare homozygous frameshift variant in *NRAP* in a DCM patient born of consanguineous parents. Interestingly, the *FHOD3* frameshift deletion at chr18:36652786 observed in our cohort was also found in a case from the 100,000 Genomes Project (**Supplementary Table 6**). The distribution of LoF variants in *NRAP* is displayed in **Figure 3a** and *FHOD3* in **Figure 3b** for the discovery cohort, 100,000 Genomes Project replication cohort, and gnomAD. Using LV myocardium from the patient with the *NRAP* variant, we confirmed that *NRAP* mRNA expression (using RNAseq and targeted qRT-PCR) and protein expression (on Western blot) were significantly downregulated in the patient harboring the variant compared to other CMP patients who did not harbor this variant (**Figure 3c**).

**Figure 3:**
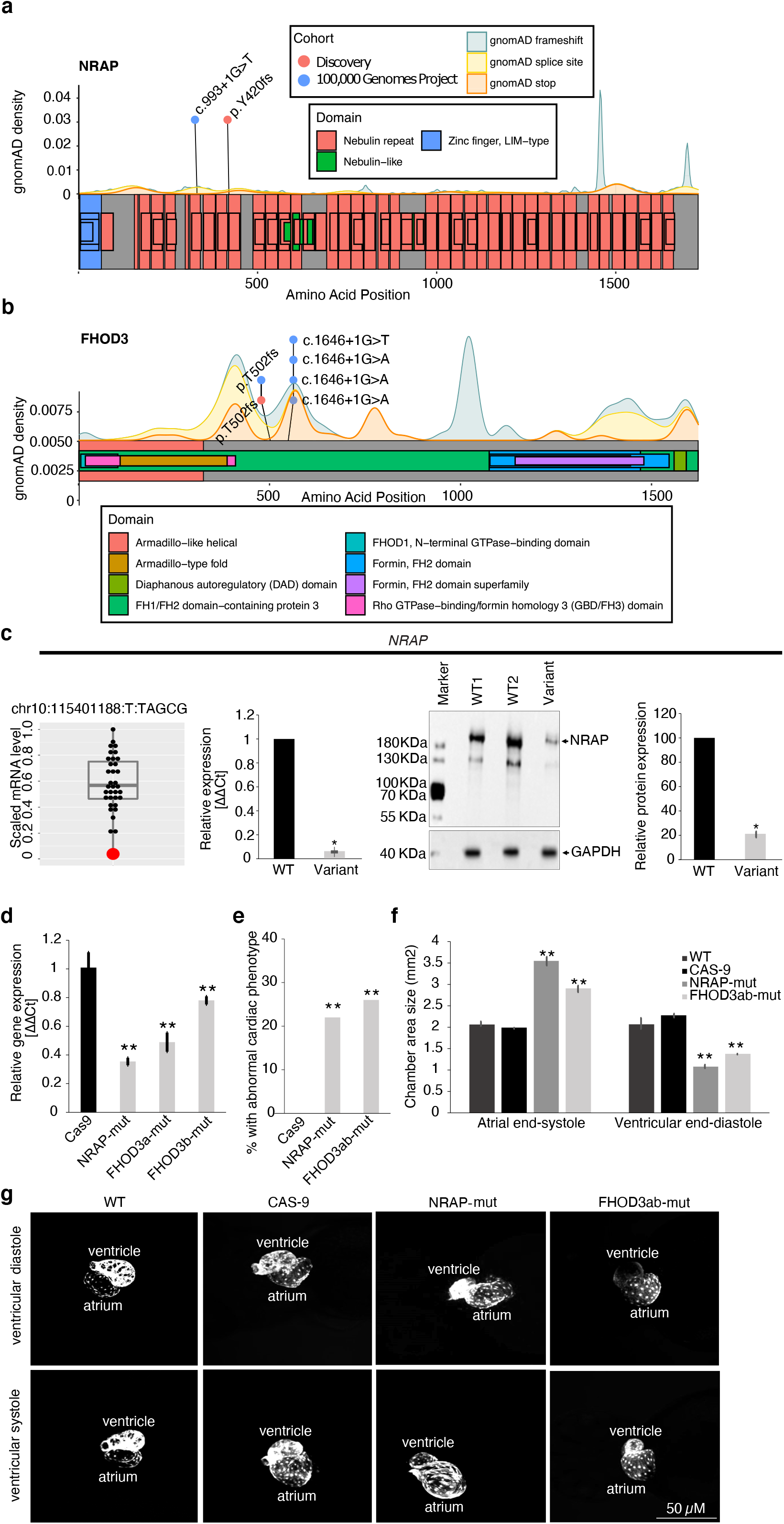
Loss of function variants in novel CMP genes: Location of loss of function variants in (**a**) *NRAP* (ENST00000359988), and (**b**) *FHOD3* (ENST00000590592) in 228 CMP cases in the discovery cohort (orange dots), and in 1326 CMP cases in the the 100,000 Genomes Project replication cohort (blue dots). gnomAD background density map of variants is shown in grey. (**c**) Myocardial *NRAP* expression: RNA-seq analysis demonstrated low *NRAP* mRNA expression (<75^th^ percentile) in the LV myocardium of a DCM patient harboring a homozygous frameshift variant (chr10:115401188_T/TAGCG) (red dot) compared to 34 CMP patients without the variant (black dots). The boxplot shows median expression for the cohort, 25^th^ and 75^th^ percentiles, and lower and upper limit values. qRT-PCR confirmed reduction of *NRAP* mRNA expression in patients with the variant compared to 2 CMP patients without the variant i.e. WT (*p<0.05 vs WT). Western blot confirmed downregulation of *NRAP* protein expression in the patient with the variant compared to 3 CMP patients without the variant on representative Western blot images (*p<0.05 vs WT). (**d-g**) Zebrafish embryos Zebrafish embryos at 1-cell stage were injected with 4 CRISPR-Cas9 guide RNA complexes to induce knockout of 2 genes, *nrap* and *fhod3ab*. (**d**) qRT-PCR showed a 35-49% downregulation of target mRNA expression in pooled samples of *nrap* and *fhod3ab* mutants compared to WT controls and Cas9 only controls (n=3 independent replicates per gene) (**p<0.01 versus controls). (**e**) 22% *nrap* mutants and 26% *fhod3ab* mutants showed an abnormal cardiac phenotype compared to 0% in Cas9 only controls (**p<0.01 vs controls). (**f**) Atrial end-systolic area was higher and ventricular end-diastolic area was lower in *nrap* and *fhod3ab* mutants compared to WT and Cas9 only controls (**p<0.01 versus controls). (**g**) Representative phase contrast images of transgenic *myl7*:GFP embryos showing cardiomegaly, atrial dilation and ventricular restriction in mutant embryos compared to wild type or Cas9 only controls. (**p<0.01 versus controls). Scale bar = 50 µm. Data are shown as mean ± standard deviations of three independent experiments per sample, with each experiment including 3 technical replicates. gnomAD, Genome Aggregation Database; WT, wild-type; mut, mutant

#### Effect of CRISPR-Cas9 knockout of novel genes in zebrafish

To confirm a role for these novel genes in cardiac structure and function *in vivo*, we induced directed knockout of *nrap* and *fhod3* in zebrafish embryos through yolk sac injection of sets of 4 CRISPR-Cas9 guide RNA (gRNA) complexes that redundantly target a single gene and induce efficient knockout to permit rapid screening for gene function^44,45^ (**Figure 3d-g)**. Sanger sequencing revealed a high burden of variants with a high cutting efficiency by 4 gRNAs targeting the exons of *nrap, fhod3a*, and *fhod3b* compared to 0% in Cas9 only controls (**Supplementary Figure 1)**. qRT-PCR showed a 0.64-fold downregulation of *nrap*, and a 0.4-fold downregulation of *fhod3a* and *fhod3b* in CRISPR-Cas9 edited embryos compared to controls (**Figure 3d**). Phenotypic analysis revealed significant atrial enlargement in gene-edited embryos compared to either wild type or Cas9 only controls (p<0.01 vs controls for all genes) (**Figure 3e**). Ventricular end-diastolic area was significantly reduced in gene-edited embryos compared to wild-type or Cas9 controls (**Figure 3f-g**) but ventricular ejection fraction was preserved (wild type 36±2%, Cas9 34±4%, *nrap* mutants 37±3% and *fhod3ab* mutants 42±2%), suggesting a restrictive CMP phenotype in embryos with defects in *nrap*, and *fhod3*. Together, these studies provide support for a role for LoF variants in novel genes like *NRAP* and *FHOD3* in causing CMP.

### Regulatory variants of CMP genes

We generated an atlas of functionally active regulatory elements of cardiac-expressed genes across the genome. This was done by mapping non-coding regions in the human heart that putatively regulate the transcription of cardiac-active genes based on experimental data assembled from heart-related epigenetic, DNAse, and histone ChIP-seq data deposited in ENCODE and other databases^46–49^. We defined promoter regions of the CMP genes by merging the DNase-seq peaks of open chromatin and histone marks specific for promoters and enhancers in cardiac tissues. Where this information was not available, we defined promoter regions as 1.5kb upstream and 1kb downstream of the transcription start site (TSS). For this study, we focussed on promoters and enhancers of known CMP genes rather than the entire genome to avoid false-positive results related to genes with an unclear association with CMP. We mapped SNVs to the active regulatory regions and defined them as Tier 1 if they were rare i.e. MAF <0.01% in population controls, and were predicted to alter transcription factor (TF) binding by at least 3 of 4 prediction tools that predict if a sequence alteration affects a likely TFBS or chromatin effects with single-nucleotide sensitivity^50–53^ (see Methods).

We further prioritized variants that had at least a 1.3-fold enrichment in cases compared to controls, that were in regulatory elements active in the human left ventricle (LV), and that were seen in gene-elusive cases (i.e. those without pathogenic coding variants in CMP genes). This provided a final prioritized list of 52 high-risk Tier 1 variants in 19% of the cohort (**Figure 4a**). Two additional patients harbored high-risk CNVs in regulatory elements of *BAG3* and *TGFB3* **(Supplementary Table 4)**. For case-control burden analysis, we used WGS data from 1326 cancer patients without heart disease from the International Cancer Genome Consortium (ICGC)^54^. This confirmed an enrichment of regulatory variants in CMP genes in cases compared to controls (OR 2.14, 95% CI 1.60-2.86, p=5.26×10^−7^) (**Figure 4b**). **Supplementary Table 7** provides details of the high-risk regulatory variants. The top 4 genes significantly enriched for regulatory variants were in pathways related to (i) α-dystroglycan glycosylation important in sarcomere structure i.e. *FKTN* (OR 53.2, CI 2.9-991), and *DTNA* (OR 5.6, CI 2.5-12.5), and (ii) desmosomal signaling i.e. *DSC2* (OR 29.3, CI 1.4-611) and *DSG2* (OR 9.7, CI 1.2-74) (**Figure 4c**). None of the variants were *de novo* amongst probands with complete trio data. Additional candidate Tier 1 variants in these and other genes important in these two signaling systems are also described in the table even though they did not meet all the high-risk criteria.

**Figure 4:**
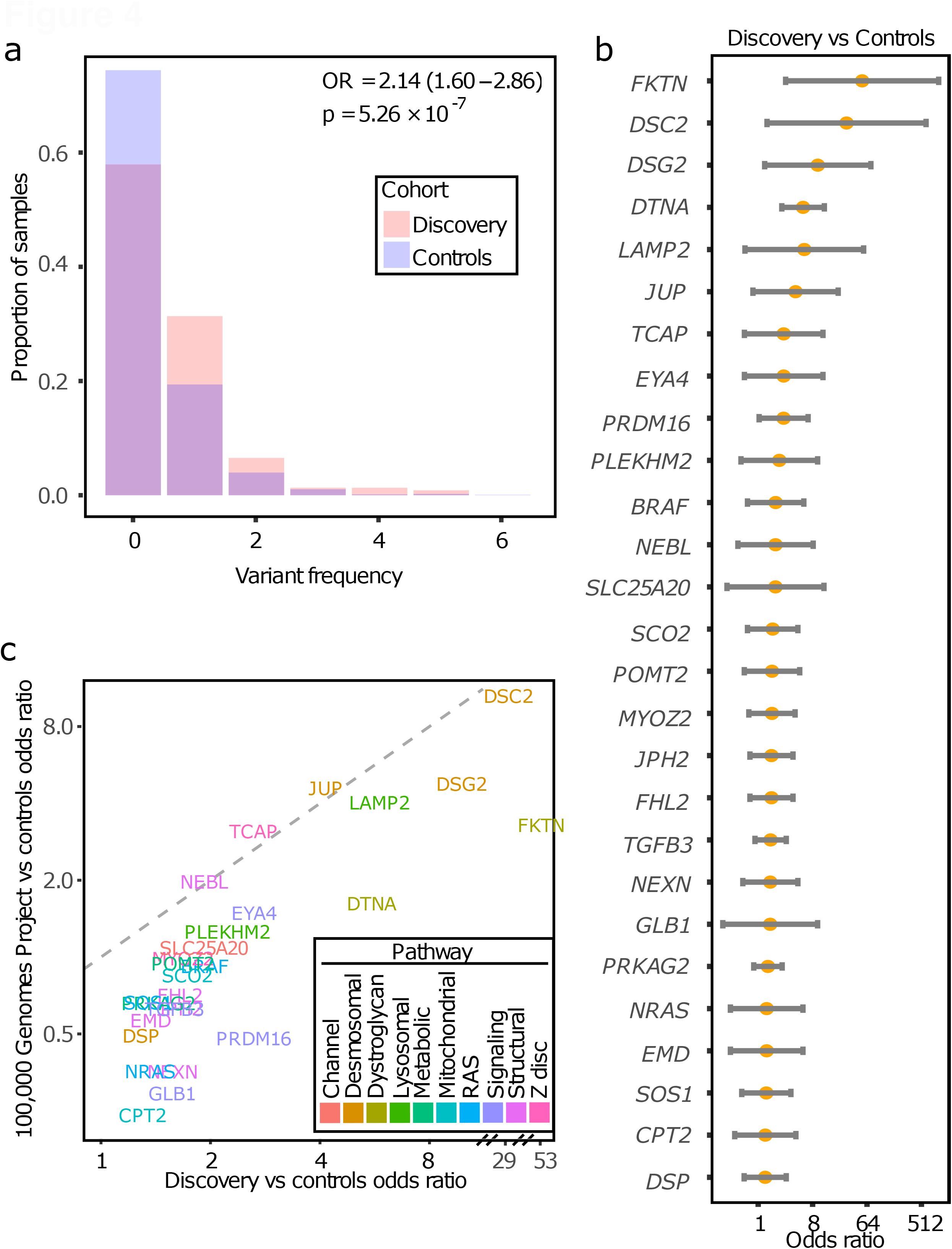
Regulatory variant burden in cases (n=228) and controls (n=1326). (**a**) Burden of Tier 1 regulatory variants in CMP genes in cases (orange) and controls (blue). There was a significant enrichment of Tier 1 regulatory variants in the cases compared to controls (OR 2.14, 95% CI 1.60-2.86, p=5.26×10^−7^). (**b**) Burden of Tier 1 regulatory variants by genes in cases in the discovery cohort versus controls. Top 4 genes enriched for regulatory variants compared to controls included *FKTN* (OR=53.2, CI=2.9-991), *DTNA* (OR=5.6, CI=2.5-12.5), *DSC2* (OR 29.3, CI 1.4-611) and *DSG2* (OR 9.7, CI 1.2-74). (**c**) Replication cohort (n=1266): Scatter plot showed positive correlation between genes enriched for high-risk regulatory variants in the CMP discovery cohort vs the 100,000 Genomes Project replication cohort (Spearman ρ^2^ 0.737, p=1.02×10^−8^) with the top genes being similar in both CMP cohorts (*FKTN, DTNA, DSC2, DSG2*) OR, Odds ratio

We extended our analysis to an independent *replication cohort* of 1266 CMP probands with WGS data from the 100,000 Genomes Project. There was a positive correlation between the discovery and replication cohorts for genes enriched for high-risk regulatory variants (Spearman ρ^2^ 0.737, p=1.02×10^−8^) with the top genes being similar in both CMP cohorts (*FKTN, DTNA, DSC2, DSG2*) with ORs ranging from 3.14-13.7 (**Figure 4d**).

#### Pathway enrichment analysis

A comparison of pathways enriched for protein-coding versus regulatory variants was performed using Gene Ontology and Reactome^55–57^ databases. Pathogenic protein-coding variants were enriched in a narrow set of gene categories directly related to muscle contraction, including binding of actin, troponin C, calmodulin, and protein kinase (**Supplementary Figure 2a**). In contrast, high-risk regulatory variants were enriched not only in genes involved in processes related to muscle contraction, but also in additional diverse pathways related to ERK/Ras signaling, fibroblast growth factor receptor signaling, and tyrosine kinase signaling (**Supplementary Figure 2b**). Unlike protein-coding variants, none of the high-risk regulatory variants were in sarcomeric genes. There were only six genes (*DSC2, DSG2, JPH2, LAMP2, NEXN, PRKAG2*) that harbored high-risk variants in both coding and regulatory regions.

Of note, a high proportion i.e. 44 (19%) cases harbored multiple coding and/or regulatory variants in known CMP genes which included 5% with multiple protein-coding variants, 4% with multiple regulatory variants, and 10% with a combination of both variant types (**Figure 1d**). Multiple variants in a third of patients were in genes important in myocardial architecture i.e. sarcomeric, cytoskeletal, desmosomal and other structural genes. Multiple variants were more common in HCM cases compared with other CMP subtypes (OR=3.4, CI=1.7-6.6, p=5.75×10^−4^).

### Functional assessment of regulatory variants

We prioritized Tier 1 regulatory variants in 6 genes (*BRAF, DSP, DTNA, FKRP, FKTN, LARGE1, PRKAG2, TGFB3)* for functional analyses based on the availability of left ventricular (LV) myocardium from variant-positive patients. **Figure 5** shows high-risk regulatory variants identified in these eight genes in our discovery cohort and the 100,000 Genomes Project cohort, overlaid on the background of the frequency distribution in the Genome Aggregation Database (gnomAD) reference population^33^. Most of the regulatory loci were depleted of variants in gnomAD suggesting highly constrained loci. **Supplementary Figure 3** shows the single nucleotide change in the variant of interest in our discovery cohort compared to wild-type sequence and the predicted effect on TF binding motifs^58^.

**Figure 5:**
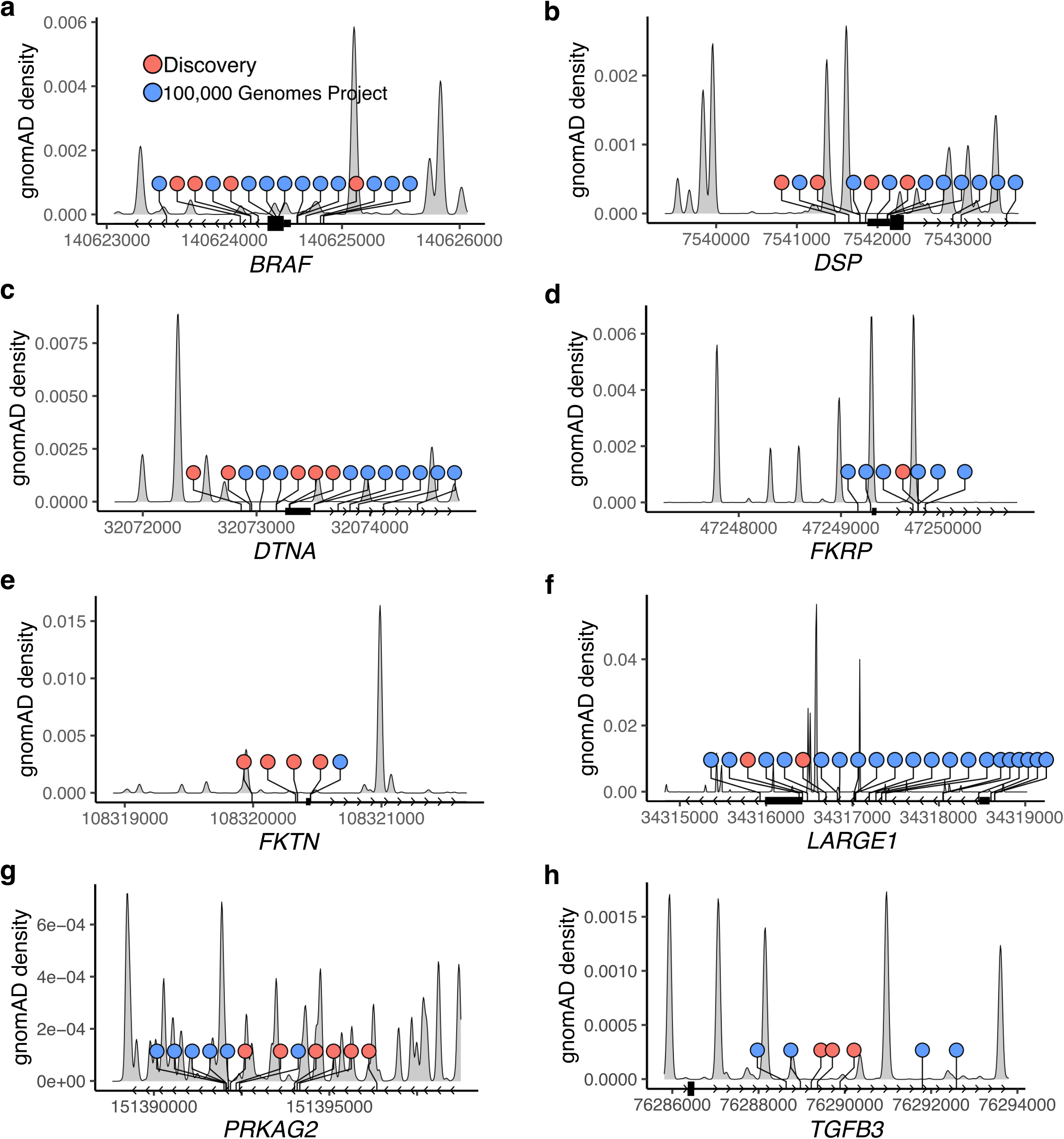
Genomic location of variants in regulatory elements of genes prioritized for functional studies. The figures show the genomic coordinates of SNVs in the discovery cohort (n=228, orange dots) and the 100,000 Genomes Project replication cohort (n=1266, blue dots) mapped relative to the first (P1) promoter region and transcription start site for the following genes (**a**) *BRAF*, (**b**) *DSP*, (**c**) *DTNA*, (**d**) *FKRP*, (**e**) *FKTN*, (**f**) *LARGE1*, and to the enhancer regions for (**g**) *PRKAG2* (E15), and (**h**) *TGFB3* (E1). SNVs observed in gnomAD reference samples are plotted as grey density curves across the region. All regulatory variants were observed with an allele frequency <0.01% in gnomAD dataset, and tended to cluster in regions that were depleted for variants in gnomAD. Coordinates are based on hg19 reference genome. gnomAD, Genome Aggregation Database

#### Association of regulatory variants with myocardial gene expression

The ability to show a change in myocardial gene expression provides critical evidence for the effect of regulatory variants on endogenous gene transcription. mRNA and protein expression was measured using RNAseq, qRT-PCR and Western blot or immunohistochemistry in 35 patients where LV myocardium was available. Myocardial expression was compared in the patient harboring the variant to controls without CMP or patients with CMP not harboring the variant.

We evaluated promoter variants in *BRAF, DSP, FKTN*, and *LARGE1* proximal to the corresponding TSS that were predicted to alter TF binding. When compared to controls and/or variant-negative CMP patients, *BRAF* mRNA showed a 0.76-fold downregulation on qRT-PCR in the patient harboring a promoter variant (chr7:140624223_G/A) (**Figure 6a**). The *DSP* variant (chr6:7541776_G/A) was associated with higher myocardial expression of *DSP* on both RNA-seq (above the 75^th^ percentile for the cohort) and on qRT-PCR (1.6-fold upregulation) (**Figure 6b**). *FKTN* promoter variant (chr9:108320330_G/A) was associated with lower *FKTN* expression in a RCM patient on RNAseq, on qRT-PCR (0.5-fold downregulation), and on Western blot (0.24-fold downregulation) (**Figure 6c**). In an HCM patient harboring a *LARGE1* promoter variant (chr22:34316416_C/T), immunohistochemistry showed reduced peri-nuclear LARGE1 protein expression in the patient compared to controls (**Figure 6d**). The *PRKAG2* enhancer variant (chr7:151392181_A/C) found in a DCM patient was associated with a 1.4-fold upregulation on qRT-PCR and a 1.5-fold upregulation on Western blot in patient myocardium (**Figure 6e**). Myocardial TGFB3 expression in a RCM patient with a high-risk enhancer variant (chr14:76289218_A/G) predicted to interact with the *TGFB3* promoter^48^ was associated with higher mRNA expression on RNA-seq, a 4.2-fold upregulation of mRNA on qRT-PCR, and 1.5-fold upregulation of TGFB3 protein on Western blot compared to controls (**Figure 6f**). These findings that are derived directly from the myocardium of patients harboring variants of interest confirmed that SNVs within key regulatory elements are associated with an important impact on functional gene products and provide important supporting evidence for variant pathogenicity.

**Figure 6:**
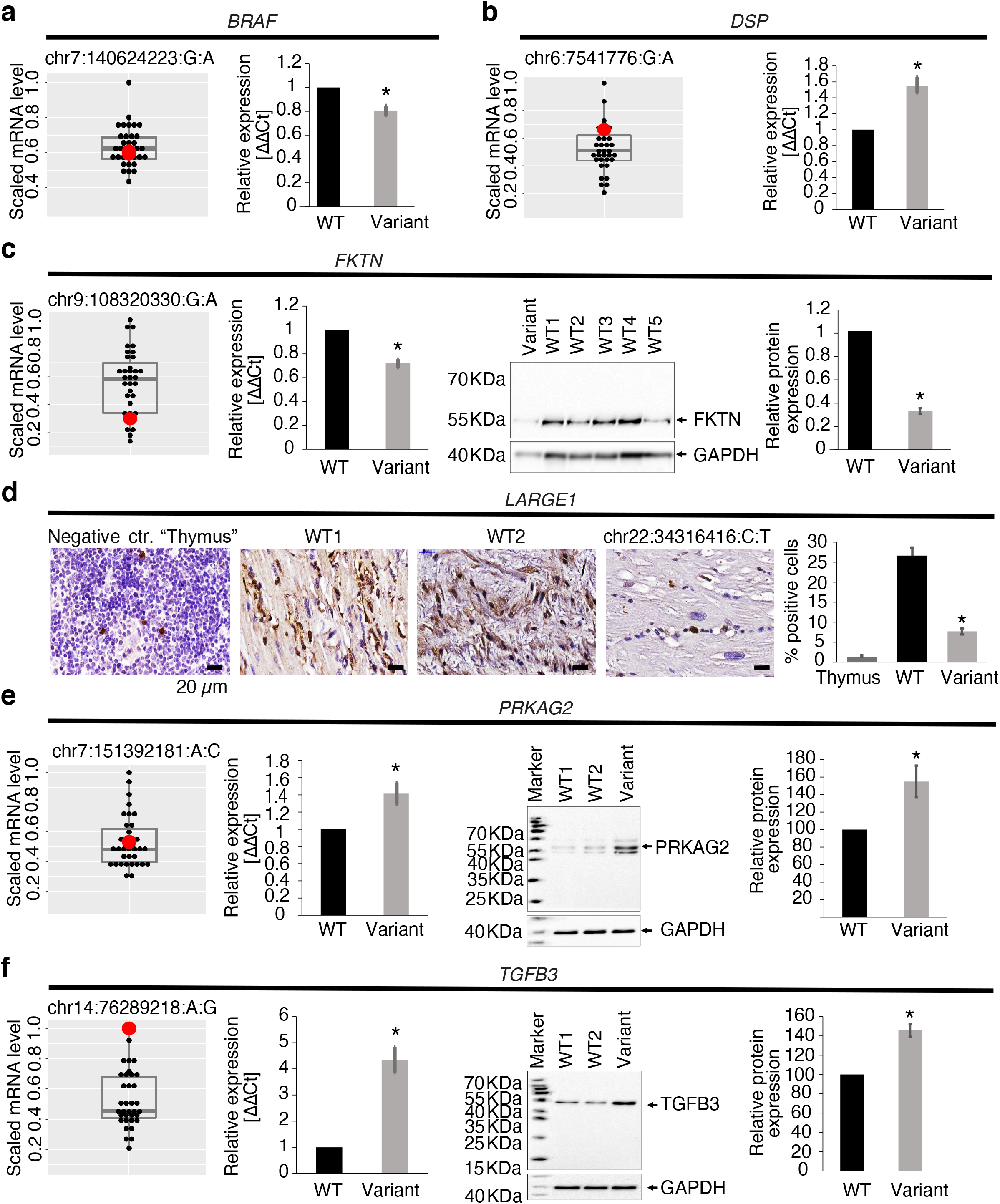
Target gene and protein expression in the LV myocardium of patients harboring regulatory variants. RNA Seq, qRT-PCR, Western blot, and immunohistochemistry were performed in available LV myocardium from CMP patients (n=35) to detect mRNA and protein expression of target genes in patients harbouring regulatory variants in *BRAF, DSP, FKTN, LARGE1, PRKAG2* or *TGFB3*. For RNA sequencing data, the target scaled RPKM gene expression was compared between the patient harboring the variant (red dot) and the remainder of the cohort (black dots) using boxplots showing median expression for the cohort, 25^th^ and 75^th^ percentiles, and maximum and minimum values (n=35). For qRT-PCR, Western blot, and immunohistochemistry, target gene or protein expression in the LV myocardium of the patient harboring the variant was compared to wild-type controls including an autopsy sample from an individual without cardiac disease as well as one or more CMP patients that did not harbour any known pathogenic coding or regulatory variants. Three independent experiments were performed per sample with each experiment including three technical replicates per sample. Protein expression level of *GAPDH* as a house keeping gene was used as a loading control for Western blots. Error bars indicate standard deviation between the averages of each independent experiment. (**a**) *BRAF*: Promoter variant chr7:140624223_G/A was associated with normal *BRAF* mRNA expression on RNAseq, but reduced *BRAF* mRNA expression on qRT-PCR. Promoter variant chr7:140624286_C/T was associated with increased mRNA expression on RNAseq (>75^th^ percentile). (**b**) *DSP*: Promoter variant (chr6:7541776_G/A) was associated with increased *DSP* mRNA expression on RNAseq (>75^th^ percentile), and on qRT-PCR (*p<0.05 vs controls). (**c**) *FKTN*: Promoter variant 1 (chr9:108320330_G/A) was associated with reduced *FKTN* mRNA expression on RNAseq (<75^th^ percentile), reduced mRNA expression on qRT-PCR (p<0.05 vs controls), reduced protein expression on Western blot representative images, and reduced relative protein abundance on quantification (*p<0.05 vs controls). (**d**) *LARGE1*: Promoter variant chr22:34316416_C/T was associated with lower perinuclear staining for LARGE1 (brown) (nuclear staining, blue) on representative immunohistochemistry images, and lower % of LARGE1 positive cells in patient myocardium (*p<0.05 vs controls). Thymic tissue was used as negative control. Scale bar = 20 µm. (**e**) *PRKAG2*: Enhancer variant chr7:151392181_A/C was associated with normal *PRKAG2* mRNA expression on RNAseq, but higher mRNA expression on qRT-PCR (*p<0.05 vs controls), higher protein expression on Western blot representative images, and higher relative protein expression on quantification (*p<0.05 vs controls). (**f**) *TGFB3*: Enhancer variant (chr14:76289218_A/G) was associated with higher *TGFB3* mRNA expression on RNAseq, higher mRNA expression on qRT-PCR (*p<0.05 vs controls), higher protein expression on Western blot representative images, and higher relative protein abundance on quantification (*p<0.05 vs controls). RNA Seq, RNA sequencing; WT, Wild-type

#### Effect of regulatory variants on gene transcription using reporter assays

##### Luciferase reporter assay

Gene promoters or enhancer+promoters harboring candidate SNVs and the corresponding control region were cloned into Firefly Luciferase reporters and transfected into human induced pluripotent stem cell (iPSC)-derived cardiomyocytes to determine the effect of the variants on the transcription activity of the luciferase reporter gene (**Supplementary Figure 4a**). The cloned promoter variants of *BRAF* (chr7:140624223_G/A), *DTNA* (chr18:32072866_A/G), *FKRP* (chr19:47249754_C/T), *FKTN* (chr9:108319991_A/C, chr9:108320330_G/A), and *LARGE1* (chr22:34316416_C/T) reduced luciferase activity compared to reference sequences. The promoter variant of *DSP (*chr6:7541776_G/A), a second promoter variant of *LARGE1 (*chr22:34316687_G/A), and an enhancer variant of *TGFB3* (chr14:76289218_A/G) significantly increased luciferase activity compared to reference sequences (**Figure 7a**). The altered transcriptional regulation of the luciferase reporter by inserting promoter and enhancer variants of several CMP genes suggests a direct regulatory effect of these SNPs on target gene transcription.

**Figure 7:**
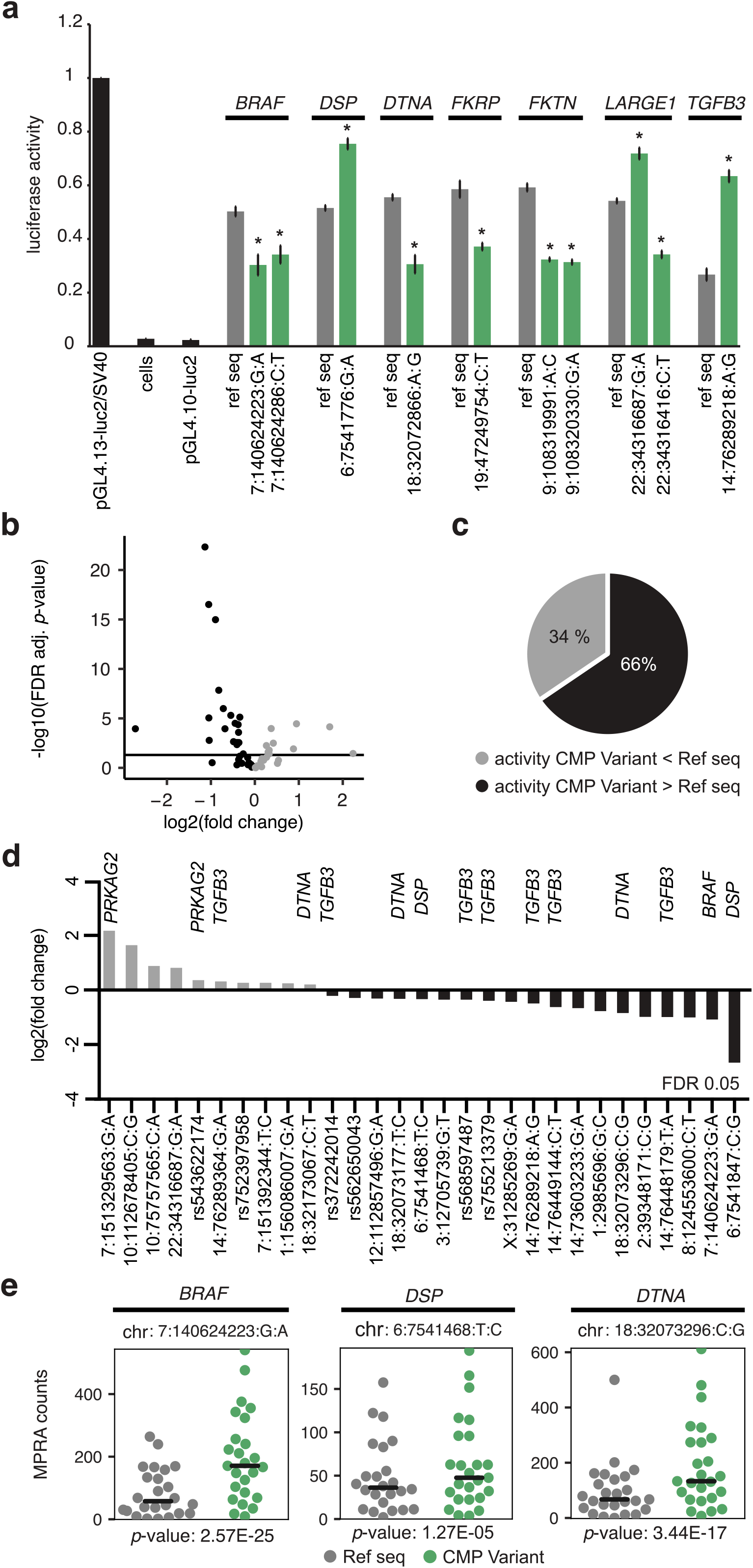
Reporter assays in human iPSC-cardiomyocytes. (**a**) Luciferase reporter assay showing the effect of regulatory variants on transcription. The cloned promoter variants of *BRAF* (chr7:140624223_G/A), *DTNA* (chr18:32072866_A/G), *FKRP* (chr19:47249754_C/T), *FKTN* (chr9:108319991_A/C, chr9:108320330_G/A), and *LARGE1* (chr22:34316416_C/T) reduced luciferase activity compared to reference sequences. The promoter variant of *DSP (*chr6:7541776_G/A), a second promoter variant of *LARGE1 (*chr22:34316687_G/A), and an enhancer variant of *TGFB3* (chr14:76289218_A/G) significantly increased luciferase activity compared to reference sequences. *p<0.05 versus reference sequence. All luciferase reporter assays were performed with 3 biological replicates, each with 3 technical replicates. (**b**) Volcano plot representing the effect of 54 regulatory variants on gene expression using MPRA. 29 variants had significant differences in transcriptional activity between reference and alternative allele (FDR<0.05, represented by horizontal black line). Grey = CMP variant activity less than reference allele; black = CMP variant activity more than reference allele. (**c**) 66% of significant variants were associated with higher transcription activity of the reference allele. (**d**) Log2-fold transcriptional activity changes between alternative and reference allele sequences. (**e**) Representative graphs of MPRA counts of alternative allele (green) versus reference allele sequences (grey) of *BRAF* (chr7:140624223_G/A), *DSP* (chr6:7541468_T/C), and *DTNA* (chr18:32073296_C/G). All MPRA assays were performed in 5 independent biological replicates. MPRA, massively parallel reporter assay; ref seq, reference allele sequence; FDR, False discovery rate; CMP, cardiomyopathy

##### Massively parallel reporter assay (MPRA)

To assess the functional effect of additional Tier 1 regulatory variants on transcriptional activity, we used the higher throughput MPRA in cardiomyocytes^22,23^. Specifically, we tested the regulatory effects of 54 variants by dissecting the transcriptional activities of their reference and alternative alleles (each allele represented by 25 unique barcodes, see Methods) in PGP17 iPSC-cardiomyocytes (n=5 independent replicates) (**Supplementary Figure 4b-e, Supplementary Table 8**). Of the 54 variants examined, 29 variants (54%) showed significant transcriptional differences between the two alleles [False discovery rate (FDR)<0.05] (**Figure 7b**,**c**) with log2-fold change ranging from -2.72 to +2.23 (**Figure 7d, Supplementary Table 8**). This represented 26 additional variants with high regulatory activity besides the ones validated in the previous myocardial and luciferase reporter assays. Representative examples for MPRA counts show high regulatory activity of variants in *BRAF, DSP*, and *DTNA* loci (**Figure 7e**). Overall, the MPRA findings confirm that our WGS confidently identified variants that are functionally active in cardiomyocytes.

In summary, our findings using WGS revealed a significant contribution of regulatory variants and CNVs in CMP genes (in 20% of cases), and a small but notable contribution of LoF protein-coding variants in novel genes (in an additional 5% of cases) to childhood onset CMP.

## DISCUSSION

WGS yields a large number of germline protein-coding and regulatory variants. An understanding of their contribution to human disease has been hampered by lack of stringent bioinformatics and functional approaches tailored to the disease under study. Using WGS in CMP, we identified deleterious protein-coding variants in 36% of our cohort, including 7.5% who had been missed despite clinical testing of candidate genes. Moreover, we found 5% of patients with deleterious variants in novel CMP genes and, very importantly, another 20% with high-risk regulatory variants not previously reported in CMP. An important subset of these regulatory variants were confirmed to have an effect on exogenous and endogenous gene expression in functional assays thereby providing strong evidence for their contribution to CMP. The discovery and validation of these novel variants reduced by half the number of gene-elusive CMP cases in our cohort.

Of the novel genes harboring deleterious protein-coding variants, two genes, *NRAP* and *FHOD3*, emerged as strong candidates. Both are important in the maintenance of sarcomeric and actin cytoskeleton in the heart and have been associated with CMP in mouse studies and small case series^36– 43,59,60^. Our proband was homozygous for a LoF variant in *NRAP* similar to a previously reported family with an autosomal recessive DCM phenotype^39^. Moreover, a high burden of variants in these genes was also found in our replication cohort. The reduced expression of *NRAP* in patient myocardium combined with the findings of reduced *nrap* and *fhod3* expression and a CMP phenotype in zebrafish knockouts provides supporting evidence for *NRAP* and *FHOD3* being novel genes that should be considered as strong candidate genes for clinical testing in CMP.

A tremendously exciting finding of our study was the enrichment of high-impact regulatory SNVs and CNVs in cases compared to controls, with 20% cases harboring these variants in recurrently mutated regions active in the human LV. When analyzed by CMP subtype, the yield of high-risk regulatory variants was higher in non-HCM CMPs in which protein-coding variants only account for a small proportion of the cases. Overall, the regulatory variants were enriched not only in pathways related to muscle contraction but also in α-dystroglycan binding, desmosomal signaling and ERK/Ras signaling. Although coding variants in these pathways typically cause multi-system involvement, we did not observe systemic features in patients harboring non-coding variants in these genes. It is possible that the effect of regulatory variants is restricted to the heart unlike coding variants that impact protein function in multiple tissues. Also, sarcomeric genes were less impacted by regulatory variants likely because they are more tolerant to haplo-insufficiency.

A notable gene set with dysregulated expression involved *DTNA, FKTN, FKRP, LARGE1*, and *POMT*, that are essential for α-dystroglycan function through post-translational glycosylation. Dystroglycan is a central component of the dystrophin-glycoprotein complex, where it functions as a transmembrane linker, anchoring the cytoskeleton to the extracellular matrix and plays a role in myocyte, sarcolemma and sarcomere stability^61,62^. Disruption of glycosylation has been associated with severe cardiac dysfunction in FKTN or LARGE1-deficient mice and with DCM (with mild to no skeletal muscle involvement) often in the context of homozygous or compound heterozygous variants^63–65^. We also found an enrichment of regulatory variants disrupting the expression of desmosomal genes (*DSG2, DSC2, JUP, DSP*) in which both missense and LoF variants have been reported to cause AVC, DCM and RCM, similar to patients in our cohort^66^.

A strength of our study was the ability to functionally validate the effect of the regulatory variants on gene and protein expression. We confirmed that the activity of a luciferase reporter gene was altered under the effect of the variant promoter/enhancer sequences compared to wild-type control in human cardiomyocytes^67,68^. We recognize that luciferase reporter assays are not able to assess for regulatory variants that affect chromatin structure. However, we defined promoters of CMP genes by merging the DNase-seq peaks of open chromatin and histone marks specific for promoters and enhancers obtained from cardiac tissue. Moreover, we were able to show that endogenous gene expression was altered in the LV myocardium of patients harboring these variants, a truly unique strength of our study. For example, upregulated myocardial TGFB3 has been observed in patients with DCM or HCM, but this is the first instance of a variant in the regulatory element of this gene being associated with upregulated *TGFB3* expression in patient myocardium^69,70^. Similarly, to the best of our knowledge, this is the first report of reduced target gene expression in the myocardium of patients harboring Tier 1 promoter variants as well as those harboring CNVs involving candidate CMP genes^71–74^. Importantly, using MPRA, we were able to demonstrate a significant regulatory effect of a larger subset of high-risk variants associated with these genes, reinforcing the strength of our variant selection strategy^75–78^. Together, these findings not only represent an important advance in our understanding of the cardiac regulome, but also provide novel insights into the genomic architecture of childhood CMP and add to the discovery of non-coding variants in human disease^21^.

Similar to previous reports, we found multiple coding variants in 5% of cases which have been reported to contribute to a more severe phenotype^79^. In our cohort, we were also able to find co-occurrence of not only coding SNVs but also CNVs. Two patients with HCM, one with a pathogenic *MYBPC3* SNV and *MYOM1* CNV died, and another with a LoF splice-site variant in *FLNC* and a deletion CNV in *CTNNA3* required heart transplantation within the first year of life. A particularly intriguing finding from our study was that as many as 14% cases harbored multiple high-risk regulatory variants, sometimes in conjunction with a pathogenic coding variant. These variants were all found in genes important in myocyte structure. Further studies are needed to examine the association of multiple regulatory variants with disease severity.

The role of regulatory variants may have been underestimated in our study since we did not explore distal enhancers. Also, TFBS that do not resemble the consensus sequence could have been misclassified as not being high-risk. As *in silico* predictions improve with time, it will enable more widespread exploration of the regulome for disease variants. Finally, we were limited in our ability to evaluate endogenous gene expression due to the small number of available myocardial samples.

Overall, our findings that high confidence variants identified using *in silico* prediction models have functional consequences validates our bioinformatics approach to novel variant discovery and makes a strong case for exploring variants in recurrently mutated cis-regulatory elements of CMP genes in order to increase the yield of genetic testing^80,81^. In summary, our work provides a guiding strategy to address regulatory variants in cardiac disease and emphasizes the need for further research to validate the clinical utility of these findings.

## METHODS

### Study cohort

The study cohort comprised unrelated primary CMP index cases less than 21 years old at diagnosis, and affected and unaffected family members, recruited between 2007-2018 through the Heart Centre Biobank at The Hospital for Sick Children, Toronto^80^. HCM, DCM, RCM, LVNC and AVC were diagnosed based on published clinical criteria^82,83^. Patients with secondary CMPs resulting from inborn errors of metabolism, mitochondrial disorders, syndromic, and neuromuscular etiologies were excluded. Clinical data including demographics, diagnosis, family history, clinical genetic testing results, and outcomes during follow-up were captured. The median age at diagnosis was 2.8 years (range 0-20), 42% were female. Major self-reported ethnicities were 71% White, 17% Asian, 6% Black. 26% of cases were genotype-positive on previous clinical panel testing, 47% were genotype-negative, and 27% were clinically untested. Ten cases (4.7%) died, and 130 cases (57%) experienced a major adverse cardiac event on follow-up (**Supplementary Table 1**). Collection and use of human DNA and myocardial tissue from CMP cases through the Heart Centre Biobank Registry was approved by the Institutional Research Ethics Boards (Hospital for Sick Children, Children’s Hospital of Eastern Ontario, Toronto General Hospital, London Health Sciences Centre, Kingston General Hospital, and Hamilton Health Sciences Centre) and written informed consent was obtained from all patients and/or their parents / legal guardians^80,81^.

### Whole genome sequencing (WGS)

WGS was performed on high quality DNA from blood or saliva to achieve a minimum of 30-fold coverage using Illumina HiSeq X platform through Macrogen, South Korea, and The Centre for Applied Genomics (TCAG, Hospital for Sick Children, Toronto). High quality paired-end reads (2×150bp) were mapped to human genome reference sequence (hg19) using Isaac aligner and variants were called using Isaac variant caller^84^. WGS quality metrics were calculated using mosdepth (https://github.com/brentp/mosdepth)^85^. Samples with average genome-wide coverage less than 10X were excluded from further analysis. Variants passing default Isaac variant caller quality metrics were annotated using snpEff (v.4.3)^86^ and annovar (v.2016.02.01)^87^. Variants used for downstream analysis were further required to have a ‘PASS’ flag in the ‘FILTER’ field. SNVs were additionally required to have a total filtered read depth (‘DP’) ≥ 10X, while indels were additionally required to have a total filtered read depth at the position preceding the indel (‘DPI’) ≥ 10X. The total number of SNVs per sample was calculated using bcftools^88^.

### Protein-coding variants in CMP genes

By mining data from Online Mendelian Inheritance in Man (OMIM) database, various commercially available CMP gene panels, manual curation from literature, we compiled a primary list of 133 candidate genes with strong association with CMPs (**Supplementary Table 2**). Mitochondrial genes were excluded. We developed computational workflows for interpretation of SNVs (missense, splicing, LoF), indels and CNVs in coding and non-coding regions.

#### Protein-coding SNVs and indels

We developed a custom variant classification workflow for the identification of pathogenic protein-coding and splice-site SNVs based on the ACMG 2015 guidelines^24^. The automated variant classification pipeline was based on information from different sources including ClinVar^89^ and Human Gene Mutation Database (HGMD)^90^ to determine previously reported pathogenic or likely pathogenic variants. 1000 Genomes^91^, NHLBI-ESP^92^, Exome Aggregation Consortium (ExAC), and genome aggregation database (gnomAD) were used as reference controls to filter for rare variants defined as MAF < 0.01%^93^. Pathogenicity of missense variants was predicted using prediction scores from at least five prediction tools including SIFT^94^, PolyPhen2^95^, MutationTaster2^96^, Mutation Assessor^97^, CADD^27^, PROVEAN^25^, phylogenetic p-value from the PHAST package for multiple alignments of 99 vertebrate genomes to the human genome (phyloP100way_vertebrate)^98^, MetaSVM and MetaLR^26^. Genomic conservation score was obtained from GERP++^99^, and phastCons^12^. Putative protein-truncating variants predicted to cause loss of function including splice-site, nonsense and frameshift variants were assessed and annotated using LOFTEE tool (https://github.com/konradjk/loftee) as a plugin via Ensembl’s Variant Effect Predictor (VEP v90) tool^100^. The pathogenicity of variants identified on clinical testing was verified using ClinVar^89^ and InterVar^101^ classifications where possible. Segregation and de novo analysis was performed on all variants when WGS from family members was available. SNVs and indels in CMP genes that met the pathogenicity criteria described above, and that further had a MAF<0.01% in the gnomAD v2.1.1 reference population, were considered causal for CMP. These likely causal variants were reviewed and confirmed through independent classification by the institutional molecular genetic testing laboratory and all reportable SNVs were confirmed using Sanger sequencing where possible.

#### Protein-coding CNVs

For CNV calling, two read-depth-based algorithms, ERDS v1.1 (estimation by read depth with SNVs)^102^ and CNVnator v0.3.2^103^, were used as previously described^29^. Identified CNV regions were annotated using a custom annotation pipeline developed at TCAG. To increase call confidence, only CNV regions >1kb in size with at least 50% reciprocal overlap between ERDS and CNVnator calls and and <70% overlap with telomeres, centromeres and segmental duplications were included in downstream analyses. Rare CNVs were defined as variants occurring at < 1% frequency in over 1500 QC pass parental samples from an autism cohort, MSSNG^18^. Using human genome CNV map^30^, CNV events overlapping CNV regions that were <30% copy number prone were prioritized for downstream analyses. Rare CNVs >1kb in size, impacting coding exons were manually inspected using reads from BAM files and were further validated using qPCR with 100% concordance. Patients that did not harbor at least one causal variant (i.e. rare, protein-coding pathogenic SNV or CNV in CMP genes) were considered gene-elusive, and were further evaluated for protein-coding variants in novel genes and in regulatory elements of known CMP genes.

#### De novo variant analysis

Complete parent-offspring trios were available in 22 cases. To identify de novo variants, we built a full Genome Analysis Toolkit (GATK)/v4.1.2.0 best practices^104^ workflow locally for joint calling of short variants (SNVs and indels) within our cohort. Paired-end raw reads were first trimmed and cleaned by trimmomatic v.0.32, then mapped to human reference genome GRCh37 per sample by using bwa v.0.7.15. The reference genome sequence and training dataset were downloaded from the GATK bundle site (ftp.broadinstitute.org/bundle/b37). Mapped reads were realigned and calibrated by Base Quality Score Recalibration (BQSR) tools. HaplotypeCaller was used to generate genotype VCF (gVCF) files for each sample. Finally, the gVCF files for all the samples were combined and joint-called by using CombineGVCFs and GenotypeGVCFs tools. In order to filter out probable artifacts in the calls, SNPs and indels were recalibrated separately by Variant Quality Score Recalibration (VQSR) tools, and variants that passed VQSR truth sensitivity level 99.5 for SNPs and level 99.0 for indels were retained. We deployed the GATK refinement workflow to identify de novo variants that were deemed pathogenic or likely pathogenic per ACMG criteria. To infer possible high confidence de novo sites, we first recalculated phred-scaled genotype likelihoods of the samples by introducing 1000 Genomes project call set (1000G_phase3_v4_20130502) and pedigrees of the trios. These additional data can be used as prior knowledge to recalibrate the confidence of the genotypes, not just calculating a sample’s genotype likelihoods only by its reads. The tool CalculateGenotypePosteriors was applied in this step. Then, we used VariantFiltration to mark out the low Genotype Quality (GQ) sites whose GQ values were lower than 20 and read depths were lower than 10. Lastly, only the sites with all trio numbers ≥ GQ 20 were defined as high confidence de novo variants in the final call set.

### Protein-coding LoF variants in novel CMP genes

To identify novel putative CMP genes beyond the 133 established CMP genes, we searched for predicted deleterious heterozygous and homozygous LoF variants (i.e. frameshift, nonsense, stopgain, stoploss, and splicing variants) in the remainder of the exome among CMP cases that did harbor a pathogenic protein-coding or high-risk regulatory variant in a CMP gene. LoF variants were identified using LOFTEE (https://github.com/konradjk/loftee)^32,33^. All LoF variants were required to be predicted as high impact by VEP^100^, observed at an allele frequency <0.01% in the gnomAD reference population, observed in <1% of unrelated families in the cohort, and affect genes that are expressed in the human heart. Variants were further prioritized if they were in a highly constrained gene (gnomAD pLI>0.9) and/or were important in heart function. Gene tissue expression level categories were obtained from the Human Protein Atlas (http://www.proteinatlas.org)^35^.

### SNVs and CNVs in regulatory elements of CMP genes

A map was generated of the regulatory regions of the human genome, primarily promoters and proximal and distal enhancers, active in the developing and adult human heart based on experimental evidence and data from the Encyclopedia of DNA Elements (ENCODE) project^47^, FANTOM project^46^, Roadmap epigenomics^49^ and published data by Dickel et al^48^. The promoter regions of all CMP genes not included by Dickel et al were defined as 1.5kb upstream and 1.25kb downstream of the transcription start site (TSS). The TSS for canonical transcripts and when necessary, cardiac transcripts in build 37 of the human genome (hg19) were downloaded from the Ensembl Genome Browser (www.ensembl.org - accessed October 2017). To identify risk SNVs within defined regulatory regions of CMP genes, an automated custom non-coding variant prioritization pipeline was developed and implemented. Briefly, variants within defined regulatory regions were annotated using Ensembl’s Variant Effect Predictor (VEP v90)^100^. Variants overlapping known Ensembl’s regulatory features were compared with those identified in WGS data in reference populations in gnomAD (n=141,456). Regulatory regions are listed in **Supplementary Table 9**. Functional impact of rare regulatory variants was assessed based on TFBS creation or disruption scores. The scores for TFBS disruption (motif loss) and TFBS creation (motif gain) were based on combined prediction scores from four different tools - RegulomeDB^50^, motifbreakR^51^, DeepSEA^52^, and Fathmm-MKL^105^. Variants were deemed Tier 1 and were scored as damaging by at least 3 of 4 prediction tools (Tier 1). Regulatory variants were further prioritized if they occurred within gene elusive CMP cases, were associated with a gene having an OR >1.3 compared to the ICGC control cohort, and were in a region that is active in the human LV. Intergenic and intronic CNVs as well as indels <1kb overlapping promoter and enhancers active in the developing and adult human heart as defined by Dickel et al^48^ were also prioritized.

### Case control regulatory variant burden analysis

WGS variant calls were obtained from 1326 patients without heart disease enrolled in the International Cancer Genome Consortium (ICGC)^54^. The WGS samples were generated from normal tissue, with 998 consisting of blood, 224 solid tissue from a site distal to the primary tumor, 76 adjacent solid tissue, and 28 other tissues. Patients included 579 females and 747 males; diagnoses included 286 pancreatic cancers, 221 brain cancers, 178 prostate cancers, 123 breast cancers, 98 esophageal cancers, 82 liver cancers, 74 renal cancers, 70 skin cancers, 68 ovarian cancers, 64 bone cancers, 37 gastric cancers, 13 oral cancers, and 12 biliary tract cancers. Data were obtained from the ICGC Data Portal Pan-Cancer Analysis of Whole Genomes (PCAWG) section. Samples were aligned to hs37d5 (GRCh37), and germline variant calls were made using the DKFZ/EMBL variant call pipeline. The “NORMAL” sample calls were extracted and filtered in a comparable way to the discovery cohort: only variants with a ‘PASS’ flag covered by at least 10 reads (DP/DPI ≥10) were used for downstream analysis. Variant calls were converted to hg19 using Picard LiftoverVcf (http://broadinstitute.github.io/picard/).

To compare variant burden between cases and controls for Tier 1 variants in regulatory elements of CMP genes, variant calls were required to have an allele frequency ≤0.01% in gnomAD. Variants observed in ≥1% of samples in the study cohort were excluded from burden testing to reduce false-positive variant calls. For each comparison (gene, pathway, or the entire regulome), ORs were calculated as the frequency of cases versus controls harboring at least one variant. P-values were calculated using a two-sided Fisher’s exact test. A false discovery rate (FDR) threshold of 0.2 was applied after removing tests where no variants were observed in the combined case and control samples. To reduce bias in these calculations and avoid “zero cells” in the contingency tables, 0.5 was added to each observed frequency (Haldane-Anscombe correction). All statistical analyses were done using R statistical software version 3.5.1.

### Replication cohort analysis

Regulatory variant burden analysis was extended to an independent cohort of 1266 CMP cases, using samples from the 100,000 Genomes Project available to us through the Genomics England Clinical Interpretation Partnership from version 8 of the main programme^106^. All cases were required to be probands with WGS data available and have at least one normalized specific disease term matching “cardiomyopathy”. Individuals with additional syndromic Human Phenotype Ontology (HPO) terms were excluded. The cohort included 745 HCM, 355 DCM, 43 LVNC, and 119 AVC subtypes; 22% were less than 21 years old at the time of diagnosis; 62% were male, 82% were of European ancestry. Where possible, short variant calls (SNVs and indels) were obtained after alignment to the reference genome hg38, otherwise GRCh37 variant calls were used. Variants were filtered to require a ‘PASS’ flag and to have a minimum total read depth (DP/DPI) of 10. hg38 and GRCh37 variant calls were converted to hg19 using Picard LiftoverVcf (http://broadinstitute.github.io/picard/). Variant burden analysis in the cases from the 100,000 Genome Project was performed as previously described by comparing with the ICGC control cohort.

### Pathway enrichment analysis

Pathway enrichment analysis was performed using g:Profiler with default parameters (https://biit.cs.ut.ee/gprofiler)^107^. The protein-coding gene set was ranked according to the total number of pathogenic SNVs, indels, and CNVs observed in our cohort. The regulatory gene set was ranked according to the total number of prioritized regulatory variants observed among gene-elusive cases. Adjusted p-values were calculated using a Bonferroni correction, and only pathways with an adjusted p-value <0.05 were considered significant.

### Subgroup analyses

To compare variant burden in protein-coding CMP genes or pathways between CMP subtypes, a series of 2×2 contingency tables were constructed whereby each subtype was tested against all others for each gene or pathway. A case was considered positive if it harbored at least one pathogenic variant (SNV, indel, and/or CNV), otherwise it was considered negative. Similarly, tests for associations with clinical outcome utilized 2×2 contingency tables and a case was considered positive if it harbored at least one variant of interest. Equivalently, burden tests for multiple variants of any type labeled ‘positive’ cases as those that harbor two or more of any variants for the gene or pathway being tested. P-values were calculated using a two-sided Fisher’s exact test. To reduce bias in the OR calculations and avoid “zero cells” in the contingency tables, 0.5 was added to each observed frequency (Haldane-Anscombe correction). A false discovery rate (FDR) was applied after removing tests where no variants were observed in any samples for each test set (genes or pathways). To identify enrichment for sarcomeric/cytoskeletal genes among all prioritized regulatory variants, a two-sided binomial test was used. Each variant was considered a ‘success’ if the variant was associated with a sarcomeric gene and was considered a ‘failure’ if the variant was associated with a different gene category. The prior probability of ‘success’ was set at 8/133 i.e. equal to the fraction of sarcomeric genes among the total set of known CMP genes. All statistical analyses were done using R statistical software version 3.5.1.

### Myocardial gene and protein expression

LV myocardium was obtained from CMP patients who had consented to biobanking from leftover tissue at the time of cardiac surgery or cardiac transplantation and was immediately snap frozen in the operating room and stored in liquid nitrogen.

#### RNA sequencing (RNAseq)

To measure myocardial gene expression, RNAseq was performed using Illumina HiSeq 2500 platform at TCAG in 35 LV samples. Total RNA was extracted from LV myocardial samples using the RNeasy Mini kit (QIAGEN, Canada). The generated raw sequence data was filtered according to the procedures described previously^108^. The filtered sequence reads were aligned to the human genome browser UCSC hg19, using Tophat v.2.0.11, and processed to extract raw read counts for genes using htseq-count v.0.6.1p2. Sequencing data was mapped to the human transcriptome using HISAT2 spliced aligner^109^, and gene expression level was quantified using StringTie^110^. Reads per kilobase of transcript per million generated were normalized for the size of each library, and normalized for the length of the transcripts. Normalized RNAseq data for the genes analyzed in this study are available in **Supplementary Table 10**. Expression analysis was performed to determine fold-difference in mRNA expression in the variant-positive patient compared to the average values in the remaining cohort (i.e. patients without the candidate SNV or CNV on WGS)^111^.

#### qRT-PCR

For additional confirmation of a difference in the mRNA expression level of the gene harboring the variant compared to the wild type sequences, we determined the relative mRNA expression using qRT-PCR^112^. Total RNA was extracted from patient LV myocardium using mirVana™ PARIS™ RNA and native protein purification Kit (Invitrogen, Carlsbad, California, USA) following the manufacturer’s protocol. The concentration and purity of the RNA was assessed using a Nanodrop 2000c (Thermo Fisher, Waltham, Massachusetts, USA). RNA with an A260/280 ratio of 2.0±0.05 was further evaluated for its integrity using a TapeStation 4200 (Agilent, Santa Clara, California, USA). RNA samples with RNA Integrity number above 5 and rRNA ratio of 1.7-2.0 were used to synthesize complementary DNA (cDNA) using SuperScript IV Reverse Transcriptase (Invitrogen, Carlsbad, California, USA). Specific oligonucleotide primers for each variant (**Supplementary Table 11**) were designed by primer3-NCBI (*https://www.ncbi.nlm.nih.gov/tools/primer-blast/*), and synthesised by Integrated DNA technologies (Coralville, Iowa, USA). Glyceraldehyde-3-phosphate dehydrogenase (GAPDH, human) was used as a housekeeping gene for normalization. The qRT-PCR was performed in a ViiA7 qPCR system (Applied Biosystems, Foster City, California, USA) using PowerUp SYBR^™^ Green Master Mix (Applied Biosystems, Foster City, California, USA). The total volume of the PCR reaction was 10 μl and PCR conditions consisted of a hold stage of 50°C for 2 min, then 95°C for 2 min followed by 40 cycles of 15 sec at 95°C and 15 sec at 55-60°C (Primer Tm dependent) and 72°C for 1 min. The relative quantification of mRNA was performed using the 2^−ΔΔ*C*t^ method^113^. mRNA expression of target genes in the LV myocardium of the patient harboring the variant was compared to wild-type tissues derived from other individuals, including an autopsy sample from an individual without cardiac disease as well as CMP patients that did not harbour any known pathogenic coding or regulatory variants. Experiments were performed three independent times and each experiment included 3 technical replicates. Differences between patients with and patients without the variant were analyzed using the Student’s unpaired t test and considered significant at p<0.05.

#### Western blot

To determine if change in mRNA expression was associated with a change in protein expression, Western blots were performed to assess myocardial protein expression^114,115^. Frozen tissues were homogenized in liquid nitrogen and lysed in radio-immunoprecipitation assay (RIPA) buffer and a protease inhibitor cocktail (Sigma, St. Louis, MO, USA). Samples were mixed with loading buffer, heated at 90°C for 5 min, separated using SDS-blot 4-12% Bis-Tris plus (Invitrogen, Carlsbad, California, USA) and transferred to nitrocellulose membrane. After blocking the membrane with 5% non-fat dry milk in phosphate buffer saline (PBS; pH:7.4), the membrane was incubated with either *FKTN* rabbit monoclonal antibody (ab131280; abcam, Cambridge, UK), rabbit polyclonal *TGF*β*3* antibody (ab15537, abcam, Cambridge, UK), rabbit *PRKAG2* Polyclonal antibody (MBS9134285, MyBiosource, San Diego, California, USA) or *NRAP* polyclonal antibody (PAS-88772; Invitrogen, Carlsbad, California, USA) in blocking buffer for 2 hour (h) at room temperature (**Supplementary Table 12**). The reference gene *GAPDH* (ab8245, abcam, Cambridge, UK) was used as a loading control. After extensive washing of the membrane with PBS/Tween-20, the goat anti-rabbit IgG-HRP and goat anti-mouse IgG-HRP (Invitrogen, Carlsbad, California,) were used as secondary antibodies at a dilution 1:2000 for 1 h at room temperature. Reactive bands were visualized by ChemiDoc MP imaging system (Bio-Rad, Hercules, California, USA). Protein expression in the LV myocardium of the patient harboring the variant was compared to control samples of other CMP patients who did not harbor this variant. The results were quantified using ImageJ software (*http://rsb.info.nih.gov/ij/*) and relative protein abundance of the immunoblot signal from each target protein was normalized to average abundance of the immunoblot signal of control samples. Statistical analyses were performed using the Student’s unpaired t test on data from 2 independent experiments. Differences were considered statistically significant at p<0.05.

#### Immunohistochemistry (IHC)

Formalin-Fixed Paraffin-Embedded (FFPE) LV tissue from a CMP patient with a *LARGE1* promoter variant and controls without *LARGE1* variants were used for IHC analysis using standard techniques^116^. FFPE tissue blocks were sectioned at 4 μm, dewaxed in xylene, dehydrated with a serial dilution of ethanol solution and washed with PBS. Antigen retrieval was performed in target retrieval solution (Dako, Burlington, ON, Canada) for 45 min followed by blocking of tissues in 3% hydrogen peroxidase (H_2_O_2_) for 10 minutes. After washing with PBS, tissue sections were incubated for 30 min at room temperature with primary antibody for anti-LARGE1 (PA5-78393, Thermo Fisher, Waltham, Massachusetts, USA) followed by incubation of sections with biotinylated secondary antibody for another 30 min (**Supplementary Table 12)**. Immunolabeling was detected using EnVision+ System-HRP DAB kits (Dako, Burlington, ON, Canada). Sections were examined and imaged with a light microscope. Cell nuclei were counterstained with Myer’s Hematoxylin Histological Staining Reagent (Dako, Burlington, ON, Canada). The photographs were analyzed with automated image analysis software (Image J, National Institutes of Health, Bethesda, Maryland). The number of LARGE1 positive cells was averaged in 10 fields per section and repeated in 3 replicates. Staining was compared between the individual harboring the *LARGE1* variant and the controls. Student’s unpaired t-test was used to determine differences between groups. A p-value of <0.05 was considered significant.

### Reporter assays in human iPSC-derived cardiomyocytes

#### Generation of human iPSC-cardiomyocytes

Induced pluripotent stem cells (iPSC) derived from peripheral blood lymphocytes of a healthy adult donor (PGP17), were differentiated into cardiomyocytes (CMs) using a STEMdiff Cardiomyocyte Differentiation Kit. The PGP17_11 iPSC line is devoid of any known cardiac disease variants and the protocol for differentiation into cardiomyocytes has been previously described^114^. The beating of differentiated iPSC-derived cardiomyocytes was observed at day 8 post differentiation. Cells were re-seeded at day 16 into 12-well plates for transient transfection. Cardiomyocytes were co-transfected with luciferase constructs at day 21. Transfected cells were harvested 48 h after transfection and firefly and renilla luciferase activity was measured using the Dual-Luciferase® Reporter Assay System.

#### Luciferase reporter assay

For functional validation of variant effect on gene transcription, Dual-Luciferase® Reporter Assay System (Promega, Madison, Wisconsin, USA) was used to test and compare the transcription activity of a luciferase reporter gene under the effect of the variant promoter or enhancer+promoter sequence from the patient, or genome reference sequence of each regulatory region as a wild-type control^67,68^. In order to generate the luciferase plasmids harboring the sequence of regulatory element of the predicted variants and wild-type as a control, the nucleotide sequences of 1.5-Kb of promoter region of *BRAF, DSP, DTNA, FKRP, FKTN* and *LARGE1*, and 2-Kb of-enhancer/promoter region of *TGFB3*, containing the strongest transcriptional activation region, were commercially synthesized (**Supplementary Table 13**) (Synbio Technologies, Monmouth Junction, NJ, USA). The commercial plasmids encoding the respective wild-type, enhancer or promoter variant sequences were digested with appropriate restriction enzymes and cloned separately into multiple cloning sites of Firefly Luciferase basic vectors (pGL4.10-luc2; Promega, Madison, Wisconsin, USA). Human iPSC-derived cardiomyocytes were seeded in 12-well plates, and co-transfected with 2 µg firefly luciferase vectors (pGL4.10-luc2; Promega, Madison, Wisconsin, USA) harboring regulatory sequences of wild type, *BRAF, DSP, DTNA, FKRP, FKTN* and *LARGE1* or *TGFB3* variants and 40 ng of Renilla Luciferase control reporter vectors (pRL-TK Vector; Promega, Madison, Wisconsin, USA) for normalization of transfection conditions. At 48 h post-transfection, luminescence was detected with Dual-Luciferase® Reporter (DLR™) assay system. The experiment was performed in three independent replicates and each sample was also tested in triplicate in each experiment. Firefly luciferase was measured, and followed by Renilla luciferase, in the same well. The normalizing activity of the experimental reporter was calculated by dividing the firefly luciferase signal by the internal renilla luciferase signal. Promoter-driven control firefly luciferase vector (pGL4.13-luc2/SV40; Promega, Madison, WI, USA) was used as a reference. An unpaired two-tailed Student’s t-test was used to compare if the difference between luciferase activity of the luciferase reporter gene under the effect of the regulatory variant sequence and reference sequence of each regulatory region as a wild-type control. The significance threshold was set at p<0.05.

#### Massively parallel reporter assay (MPRA)

Oligonucleotides of 135 bp with 11-bp barcodes were designed and synthesized by TwistBioscience (USA). Variants were centered within the 135 bp oligo. The full list of variants tested can be found in **Supplementary Table 6**. To control for technical variation and to assess biological relevance, each tested allele was represented a minimum of 25 times, each with a unique barcode. The oligonucleotide library contained 2700 oligos for our genomic variants, 100 oligonucleotides for positive controls, and 1500 oligonucleotides for negative controls i.e. scrambled sequences. These oligonucleotides were part of an oligonucleotide library that included an additional 234,500 sequences as part of a larger study. The cloning strategy of the oligonucleotide library and selection of positive negative controls (300 random sequences, each with 5 barcodes) was performed according to Mattioli et. al^23^. The oligonucleotide library was transfected into five biological replicates of PGP17 iPSC-derived cardiomyocytes with over 80% transfection efficiency across all replicates, using Lipofectamine Stem Transfection Reagent (STEM00015 Thermo Fisher, Waltham, Massachusetts, USA) (**Supplementary Figure 3b**). 48 hours post transfection, total RNA was harvested and DNA contamination was removed using DNase I (18047019, Thermo Fisher, Waltham, Massachusetts, USA). RNA samples with RNA Integrity number >7 were used to synthesize cDNA using SuperScript IV Reverse Transcriptase (Invitrogen, Carlsbad, California, USA). cDNA was used for library synthesis if it lacked plasmid contamination as determined by qRT-PCR performed on a ViiA7 qPCR system (Applied Biosystems, Foster City, California, USA) using PowerUp SYBR^™^ Green Master Mix (Applied Biosystems, Foster City, California, USA) (**Supplementary Figure 3c**). Tag-seq libraries were prepared as previously described^23^, and sequenced with single-end 50 bp reads on the HiSeq2500 platform (TCAG, Hospital for Sick Children, Toronto). Data were analyzed using MPRAAnalyze software ^23,117^ using random oligonucleotide sequences as null distribution. P values were calculated using a likelihood ratio test with MPRAAnalyze and a FDR<0.05 was considered significant.

### CRISPR-Cas9 editing to evaluate novel gene function in zebrafish embryos

All zebrafish embryo studies were performed at the SickKids Genetics and Disease Models Core (Zebrafish Core), Toronto, and approved by the SickKids Animal Care Committee (Protocol #401951).

#### Guide RNA (gRNA) design, synthesis and microinjection

All gRNA sequences were adapted from^45^, and are described in **Supplementary Table 14**. The primer sequences (**Supplementary Table 15**) were synthesized by Integrated DNA technologies (IDT, Coralville, Iowa, USA) and used for sgRNA *in vitro* synthesis, according to the earlier described protocol^45^. Microinjections were performed as described previously^45^ with minor modifications. Briefly, for *nrap* gRNA1, 250 pg of each gRNA with 800 pg Cas9 protein (Alt-R^®^ S.p. Cas9 Nuclease V3, cat #1081058, IDT, Coralville, Iowa, USA) were co-injected into wild-type embryos at one cell stage. For the co-injection of 8 gRNAs of *fhod3a+b*, gRNA1-gRNA4, 125 pg of each gRNA was injected while the amount of Cas9 protein remained unchanged. The injected embryos were kept in 0.003% Phenylthiourea (PTU) solution and incubated in a dark incubator at 28.5 °C for 3 days. All phenotypic analysis, imaging, DNA extraction and sequencing were performed at 3-days post fertilization (dpf).

#### DNA extraction, PCR and sequence analysis

Crude DNA was extracted from whole zebrafish larvae using 1X-PCR buffer (10 mM KCl, 10 mM Tris, PH 8.0; 1.5 mM MgCl2) containing 1 mg/ml proteinase K (Thermo Scientific, Waltham, Massachusetts, USA). The mixture was incubated at 55 °C for 50 min and then 98 °C for 10 min to deactivate proteinase K. To sequence each gRNA region, PCR was performed using Taq DNA polymerase (Bio basic, Markham, ON, Canada). The 25 μl reaction mixture contained 1X-PCR reaction buffer, 2 mM MgCl2, 0.2 mM dNTP, 0.2 mM of each forward and reverse primers, 0.75 U of Taq polymerase, and 1.5 μl of crude DNA (∼200 ng). The primer pairs and their corresponding annealing temperatures are summarized in **Supplementary Table 15**. The PCR reactions were set up as follows: 95 °C for 5 min, followed by 35 cycles of 95 °C for 20s, anneal temperature for 1 min, 72°C for 1 min and the final elongation is 72 °C for 5 min. The PCR product was purified using ExoSAP-IT (Applied Biosystems, Foster City, California, USA) following the manufacturer’s instructions and 100 ng of each PCR product was sent for sequencing to TCAG (Toronto, ON, Canada) with sequencing primers described in **Supplementary Table 15**. The sequencing results were analyzed using ICE Analysis (https://ice.synthego.com/#/) or Geneious 9.1.4.

#### qRT-PCR

At 3 dpf, pooled RNA samples were collected either from zebrafish larvae injected with gRNAs of target genes or Cas9 only as a control using TRIzol™ Reagent (Invitrogen, Carlsbad, California, USA). First-strand cDNA was synthesized using high capacity cDNA reverse transcription kit (Applied Biosystems, Foster City, California, USA) following the manufacturer’s instructions. The primers listed in **Supplementary Table 16** were used to amplify two reference genes of β*–actin* and *GAPDH* to normalize data. qRT-PCR assay was performed in a Roche LightCycler 96 machine using PowerUp SYBR Green Master Mix (Applied Biosystems, Foster City, California, USA). The relative expression level was calculated based on two technical repeats using 2^-ΔΔCT^ method^113^.

#### Sequencing

DNA samples were extracted from whole zebrafish larvae at 3 dpf and submitted for Sanger sequencing to TCAG (Toronto, ON, Canada) to confirm cutting efficiency in the exons targeted by *nrap, fhod3a*, and *fhod3b* gRNA compared to Cas9 only as a control. ICE CRISPR analysis tool (Synthego, Menlo Park, CA) was used for analysis of CRISPR edits in *fhod3b*.

#### Zebrafish cardiac phenotyping

Cardiac phenotyping of zebrafish embryos was performed at 3 dpf to assess cardiac chamber morphology, size and function. For wild field microscope *in vivo* imaging, 3 dpf zebrafish larvae were anesthetized with 0.02% tricaine and mounted in 3% methylcellulose in 50 mm glass-bottomed dishes. Video imaging was done with the Zeiss AXIO Zoom V16 Microscope using a PlanNeoFluar Z 1X/0.25 FWD 56mm objective lens under 112x magnification. The Movie Recorder function under Zen pro program was used and approximately 100 frames were captured for each video. All videos were exported at 17 frames per second for further analysis. Images were captured with a Nikon Eclipse Ti microscope under the Nikon A1 plus confocal imaging system using NIS-Elements program. Atrial area was measured at end-systole, and ventricular area was measured at end-systole and end-diastole with ventricular ejection fraction defined as (end-diastolic area – ventricular systolic area) / ventricular end-systolic area x 100 using ImageJ (*https://imagej.nih.gov/ij/*). Atrial and ventricular sizes and ventricular ejection fraction were compared using a two-tailed Student’s unpaired t-test to measure significant differences between mutants (*nrap* and *fhod3*) and controls (Cas9 and wild-type). Differences were considered statistically significant at P<0.05.

#### Data availability

Sequencing data are currently being deposited in the EGA European Genome-Phenome Archive and will be available for download upon approval by the Data Access Committee. Additional data generated or analyzed during this study are included in the supplementary information files, and additional raw data used for figures and results are available from the corresponding author on reasonable request. All computational tools used in this study are available as commercial or open-source software.

## ACKNOWLEDGEMENTS

We acknowledge the Labatt Family Heart Centre Biobank at the Hospital for Sick Children for access to DNA samples for whole genome sequencing, and The Centre for Applied Genomics at the Hospital for Sick Children for performing sequencing. We thank Xiucheng Cui and Emanuela Pannia for performing the zebrafish experiments at the SickKids Zebrafish Genetics and Disease Models Core (CRISPR-Cas9 and gRNA syntheses, zebrafish embryo microinjections, gRNA PCR validation, qRT-PCR, cardiac imaging). This research was made possible through access to the data and findings generated by the 100,000 Genomes Project. The 100,000 Genomes Project uses data provided by patients and collected by the National Health Service as part of their care and support. We thank members of the ICGC/PCAWG working groups for generating the variant calls used in our case-control burden analyses.

## CONSORTIA

### Genomics England Research Consortium

Ambrose, J. C.^1^; Arumugam, P.^1^; Baple, E. L.^1^; Bleda, M.^1^; Boardman-Pretty, F.^1,2^; Boissiere, J. M.^1^; Boustred, C. R.^1^; Brittain, H.^1^; Caulfield, M. J.^1,2^; Chan, G. C.^1^; Craig, C. E. H.^1^; Daugherty, L. C.^1^; de Burca, A.^1^; Devereau, A.^1^; Elgar, G.^1,2^; Foulger, R. E.^1^; Fowler, T^1^.; Furió-Tarí, P.^1^; Giess A.^1^; Hackett, J. M.^1^; Halai, D.^1^; Hamblin, A.^1^; Henderson, S.^1,2^; Holman, J. E.^1^; Hubbard, T. J. P.^1^; Ibáñez, K.^1,2^; Jackson, R.^1^; Jones, L. J.^1,2^; Kasperaviciute, D.^1,2^; Kayikci, M.^1^; Kousathanas, A.^1^; Lahnstein, L.^1^; Lawson, K.^1^; Leigh, S. E. A.^1^; Leong, I. U. S.^1^; Lopez, F. J.^1^; Maleady-Crowe, F.^1^; Mason, J.^1^; McDonagh, E. M.^1,2^; Moutsianas, L.^1,2^; Mueller, M.^1,2^; Murugaesu, N.^1^; Need, A. C.^1,2^; Odhams, C. A.^1^; Orioli A.^1^; Patch, C.^1,2^; Perez-Gil, D.^1^; Pereira, M. B.^1^; Polychronopoulos, D.^1^; Pullinger, J.^1^; Rahim, T.^1^; Rendon, A.^1^; Riesgo-Ferreiro, P.^1^; Rogers, T.^1^; Ryten, M.^1^; Savage, K.^1^; Sawant, K.^1^; Scott, R. H.^1^; Siddiq, A.^1^; Sieghart, A.^1^; Smedley, D.^1,2^; Smith, K. R.^1,2^; Smith, S. C.^1^; Sosinsky, A.^1,2^; Spooner, W.^1^; Stevens, H. E.^1^; Stuckey, A.^1^; Sultana, R.^1^; Tanguy M.^1^; Thomas, E. R. A.^1,2^; Thompson, S. R.^1^; Tregidgo, C.^1^; Tucci, A.^1,2^; Walsh, E.^1^; Watters, S. A.^1^; Welland, M. J.^1^; Williams, E.^1^; Witkowska, K.^1,2^; Wood, S. M.^1,2^; Zarowiecki, M.^1^

^1.^ Genomics England, London, UK

^2.^ William Harvey Research Institute, Queen Mary University of London, London, EC1M 6BQ, UK.

## AUTHOR CONTRIBUTIONS

RL, AS, JE, SM conceptualized and designed the work; RL, AS, OA, JB, TL, RY, FM, RRN, AM, QY, GM, MCSY, WWL, BT, GERC, JL, EO, LB, JS, TM, JE, SWS, SM acquired, analyzed or interpreted the data; RL, OA, TL, RY, MCSY, WWL, BT performed the bioinformatics analysis; KM, KD, WO, MM, PGM designed, executed and analyzed the MPRA dataset; RL, AS, SM drafted the original manuscript; RL, AS, PGM, JE, SWS, SM substantively revised it; and all authors reviewed and approved the final manuscript.

## SUPPLEMENTARY FIGURE LEGENDS

**Supplementary Figure 1: Sequencing results of gene editing in zebrafish**. Sanger sequencing of gene-edited zebrafish (n=7) revealed a high burden of mutations in the exons targeted by 4 gRNAs for (a) *nrap*, and (b) *fhod3ab* compared to Cas9 only controls. Panels show the reference sequence, location of the gRNA, the targeted exon, effect of Cas9 only, versus CRISPR/Cas9 gRNA in zebrafish embryos. SNVs, indels and large deletions are seen in brown and blue. *nrap, fhod3a* and *fhod3b* gRNAs caused CRISPR edits in all injected embryos and were analyzed using Synthego ICE software as needed.

Ref-seq, reference sequence: gRNA, guide RNA; ICE, inference of CRISPR edits

**Supplementary Figure 2: Pathways enriched for protein-coding and regulatory variants in the overall cohort (n=228)**. (**a**) Gene Ontology (molecular function category) and Reactome pathways enriched for pathogenic protein-coding and splicing variants. (**b**) Pathways enriched for high-risk Tier 1 regulatory variants including muscle-related categories, dystroglycan binding, fibroblast growth factor receptors, and Ras pathways.

**Supplementary Figure 3: Prediction effect of regulatory variants on transcription factor binding motif**. SeqLogo was used to predict motif disruption caused by variants in the regulatory elements of (**a, b**) *BRAF*, (**c**) *DSP*, (**d**) *DTNA*, (**e**) *FKRP*, (**f, g**) *FKTN*, (**h, i**) *LARGE1*, (**j**) *PRKAG2*, and (**k**) *TGFB3*. The nucleotide base pair outlined in the red box indicates the position of the variant in the motif. Regulatory sequence analysis of variants shows a single nucleotide change in each variant compared to reference sequence resulting in a disruption in transcription factor motifs that is predicted to be associated with transcriptional up- or down-regulation of the target gene.

Major, reference sequence; Risk, variant sequence.

**Supplementary Figure 4: Luciferase assays in hiPSC-derived CMs**. (**a**) Luciferase reporter gene vectors harboring various promoter sequences were used. The promoter-driven control Firefly luciferase vector (pGL4-13_luc2_SV40) and Promoterless Firefly Luciferase Basic Vector (pGL4-10-luc2) were used as a positive and a negative control, respectively. Renilla Luciferase control reporter vectors (pRL_TK Vector) was used for normalization of transfection conditions. Sequence of regulatory elements of the predicted variants and wild-type were commercially synthesized and cloned separately into multiple cloning sites of Firefly Luciferase basic vectors, pGL4.10_luc2. hiPSC derived CMs were co-transfected with firefly luciferase vector harboring regulatory sequences separately and Renilla Luciferase control reporter vector. Luminescence was detected with Dual-Luciferase® Reporter (DLR™) assay system. (**b**) Successful differentiation (cardiac troponin T staining in red) and transfection of plasmid pX601_GFP (green) into day 21 PGP17 iPSC-derived cardiomyocytes using Lipofectamine Stem Transfection Reagent. Magnification: ×20. (**c**) qRT-PCR was performed to detect DNA contamination from plasmidpool transfection of 5 biological replicates of PGP17 cardiomyocytes. (**d**) Unimodal distribution of barcodes that represent the oligonucleotides used in this project. DNA input represents plasmid pool of oligonucleotides, whilst replicates 1-5 (each replicate split on two lanes of HiSeq2500) flowcell are tag-seq libraries derived from the transfections in cardiomyocytes. (**e**) Pearson correlation of 5 MPRA replicates. hiPSC, human induced pluripotent stem cell; CM, cardiomyocytes; GFP, green fluorescent protein; P, plasmid; Pr, promoter; En, Enhancer; Luc, Luciferase; RES, Regulatory element sequence, WT, wild-type; V, Variant; rep, replicate

## SUPPLEMENTARY TABLES

**Supplementary Table 1:** Clinical characteristics of pediatric cardiomyopathy probands in the discovery cohort (n=228).

**Supplementary Table 2:** Cardiomyopathy gene list.

**Supplementary Table 3:** Cardiomyopathy genes harboring pathogenic or likely pathogenic coding SNVs and indels (n=228 unrelated cases).

**Supplementary Table 4:** Copy number variants affecting cardiomyopathy genes (n=228 unrelated cases).

**Supplementary Table 5:** Loss of function variants in novel cardiomyopathy genes (n=228 unrelated cases).

**Supplementary Table 6**: Loss of function variants in *NRAP* and *FHOD3* in cardiomyopathy discovery (n=228) and replication (n=1266) cohorts.

**Supplementary Table 7:** High-risk (and candidate) Tier 1 SNVs in regulatory elements of cardiomyopathy genes (n=228 cases)

**Supplementary Table 8**: Tier 1 SNVs in regulatory elements of cardiomyopathy genes evaluated by MPRA (n=228 cases)

**Supplementary Table 9:** Regulatory regions of cardiomyopathy genes for mapping non-coding variants.

**Supplementary Table 10:** Normalized RNAseq data for genes with high-risk CNVs, LoF and regulatory variants

**Supplementary Table 11:** Primer pairs for qRT-PCR in LV myocardium of candidate genes harboring regulatory variants.

**Supplementary Table 12:** Antibodies used for Western blot and immunohistochemistry

**Supplementary Table 13:** Synthesis of gene promoter and enhancer sequences for luciferase assays.

**Supplementary Table 14:** Design of single guide RNAs to target novel genes in zebrafish embryos.

**Supplementary Table 15:** Primer pairs for CRISPR-Cas9 editing of novel genes in zebrafish embryos.

**Supplementary Table 16:** Primer pairs for qRT-PCR for novel genes targeted by CRISPR-Cas9 gene editing in zebrafish embryos.

## REFERENCES

1. Maron, B. J., Rowin, E. J. & Maron, M. S. Global Burden of Hypertrophic Cardiomyopathy. JACC Heart Fail. 6, 376–378 (2018).

2. Semsarian, C., Ingles, J., Maron, M. S. & Maron, B. J. New perspectives on the prevalence of hypertrophic cardiomyopathy. J. Am. Coll. Cardiol. 65, 1249–1254 (2015).

3. Lipshultz Steven E. et al. Cardiomyopathy in Children: Classification and Diagnosis: A Scientific Statement From the American Heart Association. Circulation 140, e9–e68 (2019).

4. Watkins, H., Ashrafian, H. & Redwood, C. Inherited Cardiomyopathies. http://dx.doi.org/10.1056/NEJMra0902923 https://www.nejm.org/doi/10.1056/NEJMra0902923 (2011) doi:10.1056/NEJMra0902923.

5. Jacoby, D. & McKenna, W. J. Genetics of inherited cardiomyopathy. Eur. Heart J. 33, 296–304 (2012).

6. Lafreniere-Roula, M. et al. Family screening for hypertrophic cardiomyopathy: Is it time to change practice guidelines? Eur. Heart J. 40, 3672–3681 (2019).

7. A, M. et al. A Validated Model for Sudden Cardiac Death Risk Prediction in Pediatric Hypertrophic Cardiomyopathy. Circulation vol. 142 https://pubmed.ncbi.nlm.nih.gov/32418493/ (2020).

8. Mathew, J. et al. Utility of genetics for risk stratification in pediatric hypertrophic cardiomyopathy. Clin. Genet. 93, 310–319 (2018).

9. Alfares, A. A. et al. Results of clinical genetic testing of 2,912 probands with hypertrophic cardiomyopathy: expanded panels offer limited additional sensitivity. Genet. Med. Off. J. Am. Coll. Med. Genet. 17, 880–888 (2015).

10. Ouellette, A. C. et al. Clinical genetic testing in pediatric cardiomyopathy: Is bigger better? Clin. Genet. 93, 33–40 (2018).

11. Mak, T. S. H. et al. Coverage and diagnostic yield of Whole Exome Sequencing for the Evaluation of Cases with Dilated and Hypertrophic Cardiomyopathy. Sci. Rep. 8, 10846 (2018).

12. Siepel, A. et al. Evolutionarily conserved elements in vertebrate, insect, worm, and yeast genomes. Genome Res. 15, 1034–1050 (2005).

13. Brittain, H. K., Scott, R. & Thomas, E. The rise of the genome and personalised medicine. Clin. Med. Lond. Engl. 17, 545–551 (2017).

14. Lionel, A. C. et al. Improved diagnostic yield compared with targeted gene sequencing panels suggests a role for whole-genome sequencing as a first-tier genetic test. Genet. Med. Off. J. Am. Coll. Med. Genet. 20, 435–443 (2018).

15. Minoche, A. E. et al. Genome sequencing as a first-line genetic test in familial dilated cardiomyopathy. Genet. Med. Off. J. Am. Coll. Med. Genet. 21, 650–662 (2019).

16. Michaelson, J. J. et al. Whole-genome sequencing in autism identifies hot spots for de novo germline mutation. Cell 151, 1431–1442 (2012).

17. Yuen, R. K. C. et al. Genome-wide characteristics of de novo mutations in autism. NPJ Genomic Med. 1, 160271–1602710 (2016).

18. C Yuen, R. K. et al. Whole genome sequencing resource identifies 18 new candidate genes for autism spectrum disorder. Nat. Neurosci. 20, 602–611 (2017).

19. Brandler, W. M. et al. Paternally inherited cis-regulatory structural variants are associated with autism. Science 360, 327–331 (2018).

20. Trost, B. et al. Genome-wide detection of tandem DNA repeats that are expanded in autism. Nature (2020) doi:10.1038/s41586-020-2579-z.

21. Richter, F. et al. Genomic analyses implicate noncoding de novo variants in congenital heart disease. Nat. Genet. 52, 769–777 (2020).

22. Melnikov, A. et al. Systematic dissection and optimization of inducible enhancers in human cells using a massively parallel reporter assay. Nat. Biotechnol. 30, 271–277 (2012).

23. Mattioli, K. et al. High-throughput functional analysis of lncRNA core promoters elucidates rules governing tissue specificity. Genome Res. 29, 344–355 (2019).

24. Richards, S. et al. Standards and guidelines for the interpretation of sequence variants: a joint consensus recommendation of the American College of Medical Genetics and Genomics and the Association for Molecular Pathology. Genet. Med. Off. J. Am. Coll. Med. Genet. 17, 405–424 (2015).

25. Choi, Y., Sims, G. E., Murphy, S., Miller, J. R. & Chan, A. P. Predicting the functional effect of amino acid substitutions and indels. PloS One 7, e46688 (2012).

26. Dong, C. et al. Comparison and integration of deleteriousness prediction methods for nonsynonymous SNVs in whole exome sequencing studies. Hum. Mol. Genet. 24, 2125–2137 (2015).

27. Kircher, M. et al. A general framework for estimating the relative pathogenicity of human genetic variants. Nat. Genet. 46, 310–315 (2014).

28. Jiang, Y. et al. Detection of clinically relevant genetic variants in autism spectrum disorder by whole-genome sequencing. Am. J. Hum. Genet. 93, 249–263 (2013).

29. Trost, B. et al. A Comprehensive Workflow for Read Depth-Based Identification of Copy-Number Variation from Whole-Genome Sequence Data. Am. J. Hum. Genet. 102, 142–155 (2018).

30. Zarrei, M., MacDonald, J. R., Merico, D. & Scherer, S. W. A copy number variation map of the human genome. Nat. Rev. Genet. 16, 172–183 (2015).

31. Gross, A. M. et al. Copy-number variants in clinical genome sequencing: deployment and interpretation for rare and undiagnosed disease. Genet. Med. 21, 1121–1130 (2019).

32. Cassa, C. A. et al. Estimating the selective effects of heterozygous protein-truncating variants from human exome data. Nat. Genet. 49, 806–810 (2017).

33. Karczewski, K. J. et al. The mutational constraint spectrum quantified from variation in 141,456 humans. Nature 581, 434–443 (2020).

34. Kim, M.-S. et al. A draft map of the human proteome. Nature 509, 575–581 (2014).

35. Uhlén, M. et al. Proteomics. Tissue-based map of the human proteome. Science 347, 1260419 (2015).

36. Luo, G. et al. Complete cDNA sequence and tissue localization of N-RAP, a novel nebulin-related protein of striated muscle. Cell Motil. Cytoskeleton 38, 75–90 (1997).

37. Zhang, J. Q. et al. Ultrastructural and biochemical localization of N-RAP at the interface between myofibrils and intercalated disks in the mouse heart. Biochemistry 40, 14898–14906 (2001).

38. Lu, S. et al. Cardiac-specific NRAP overexpression causes right ventricular dysfunction in mice. Exp. Cell Res. 317, 1226–1237 (2011).

39. Truszkowska, G. T. et al. Homozygous truncating mutation in NRAP gene identified by whole exome sequencing in a patient with dilated cardiomyopathy. Sci. Rep. 7, 3362 (2017).

40. Matsuyama, S. et al. Interaction between cardiac myosin-binding protein C and formin Fhod3. Proc. Natl. Acad. Sci. U. S. A. 115, E4386–E4395 (2018).

41. Wooten, E. C. et al. Formin homology 2 domain containing 3 variants associated with hypertrophic cardiomyopathy. Circ. Cardiovasc. Genet. 6, 10–18 (2013).

42. Arimura, T. et al. Dilated cardiomyopathy-associated FHOD3 variant impairs the ability to induce activation of transcription factor serum response factor. Circ. J. Off. J. Jpn. Circ. Soc. 77, 2990–2996 (2013).

43. Esslinger, U. et al. Exome-wide association study reveals novel susceptibility genes to sporadic dilated cardiomyopathy. PloS One 12, e0172995 (2017).

44. Huang, C.-J., Tu, C.-T., Hsiao, C.-D., Hsieh, F.-J. & Tsai, H.-J. Germ-line transmission of a myocardium-specific GFP transgene reveals critical regulatory elements in the cardiac myosin light chain 2 promoter of zebrafish. Dev. Dyn. Off. Publ. Am. Assoc. Anat. 228, 30–40 (2003).

45. Wu, R. S. et al. A Rapid Method for Directed Gene Knockout for Screening in G0 Zebrafish. Dev. Cell 46, 112-125.e4 (2018).

46. Andersson, R. et al. An atlas of active enhancers across human cell types and tissues. Nature 507, 455–461 (2014).

47. ENCODE Project Consortium. An integrated encyclopedia of DNA elements in the human genome. Nature 489, 57–74 (2012).

48. Dickel, D. E. et al. Genome-wide compendium and functional assessment of in vivo heart enhancers. Nat. Commun. 7, 12923 (2016).

49. Roadmap Epigenomics Consortium et al. Integrative analysis of 111 reference human epigenomes. Nature 518, 317–330 (2015).

50. Boyle, A. P. et al. Annotation of functional variation in personal genomes using RegulomeDB. Genome Res. 22, 1790–1797 (2012).

51. Coetzee, S. G., Coetzee, G. A. & Hazelett, D. J. motifbreakR: an R/Bioconductor package for predicting variant effects at transcription factor binding sites. Bioinforma. Oxf. Engl. 31, 3847–3849 (2015).

52. Shihab, H. A. et al. An integrative approach to predicting the functional effects of non-coding and coding sequence variation. Bioinforma. Oxf. Engl. 31, 1536–1543 (2015).

53. Zhou, J. & Troyanskaya, O. G. Predicting effects of noncoding variants with deep learning-based sequence model. Nat. Methods 12, 931–934 (2015).

54. International Cancer Genome Consortium et al. International network of cancer genome projects. Nature 464, 993–998 (2010).

55. Ashburner, M. et al. Gene ontology: tool for the unification of biology. The Gene Ontology Consortium. Nat. Genet. 25, 25–29 (2000).

56. The Gene Ontology Consortium. The Gene Ontology Resource: 20 years and still GOing strong. Nucleic Acids Res. 47, D330–D338 (2019).

57. Jassal, B. et al. The reactome pathway knowledgebase. Nucleic Acids Res. 48, D498–D503 (2020).

58. Schneider, T. D. & Stephens, R. M. Sequence logos: a new way to display consensus sequences. Nucleic Acids Res. 18, 6097–6100 (1990).

59. Valdés-Mas, R. et al. Mutations in filamin C cause a new form of familial hypertrophic cardiomyopathy. Nat. Commun. 5, 5326 (2014).

60. Olson, T. M. et al. Metavinculin mutations alter actin interaction in dilated cardiomyopathy. Circulation 105, 431–437 (2002).

61. Michele, D. E., Kabaeva, Z., Davis, S. L., Weiss, R. M. & Campbell, K. P. Dystroglycan matrix receptor function in cardiac myocytes is important for limiting activity-induced myocardial damage. Circ. Res. 105, 984–993 (2009).

62. Johnson, E. K. et al. Proteomic analysis reveals new cardiac-specific dystrophin-associated proteins. PloS One 7, e43515 (2012).

63. Ujihara, Y. et al. Elimination of fukutin reveals cellular and molecular pathomechanisms in muscular dystrophy-associated heart failure. Nat. Commun. 10, 5754 (2019).

64. Arimura, T. et al. Mutational analysis of fukutin gene in dilated cardiomyopathy and hypertrophic cardiomyopathy. Circ. J. Off. J. Jpn. Circ. Soc. 73, 158–161 (2009).

65. Murakami, T. et al. Fukutin gene mutations cause dilated cardiomyopathy with minimal muscle weakness. Ann. Neurol. 60, 597–602 (2006).

66. James, C. A., Syrris, P., van Tintelen, J. P. & Calkins, H. The role of genetics in cardiovascular disease: arrhythmogenic cardiomyopathy. Eur. Heart J. 41, 1393–1400 (2020).

67. Madan, N. et al. Functionalization of CD36 cardiovascular disease and expression associated variants by interdisciplinary high throughput analysis. PLoS Genet. 15, e1008287 (2019).

68. Kapoor, A. et al. Multiple SCN5A variant enhancers modulate its cardiac gene expression and the QT interval. Proc. Natl. Acad. Sci. U. S. A. 116, 10636–10645 (2019).

69. Dobaczewski, M., Chen, W. & Frangogiannis, N. G. Transforming growth factor (TGF)-β signaling in cardiac remodeling. J. Mol. Cell. Cardiol. 51, 600–606 (2011).

70. Beffagna, G. et al. Regulatory mutations in transforming growth factor-beta3 gene cause arrhythmogenic right ventricular cardiomyopathy type 1. Cardiovasc. Res. 65, 366–373 (2005).

71. Landstrom, A. P. et al. Junctophilin-2 expression silencing causes cardiocyte hypertrophy and abnormal intracellular calcium-handling. Circ. Heart Fail. 4, 214–223 (2011).

72. Li, J. et al. Loss of αT-catenin alters the hybrid adhering junctions in the heart and leads to dilated cardiomyopathy and ventricular arrhythmia following acute ischemia. J. Cell Sci. 125, 1058–1067 (2012).

73. Aherrahrou, Z. et al. Knock-out of nexilin in mice leads to dilated cardiomyopathy and endomyocardial fibroelastosis. Basic Res. Cardiol. 111, 6 (2016).

74. Prins, K. W. et al. Colchicine Depolymerizes Microtubules, Increases Junctophilin-2, and Improves Right Ventricular Function in Experimental Pulmonary Arterial Hypertension. J. Am. Heart Assoc. 6, (2017).

75. Inoue, F. & Ahituv, N. Decoding enhancers using massively parallel reporter assays. Genomics 106, 159–164 (2015).

76. Vockley, C. M. et al. Massively parallel quantification of the regulatory effects of noncoding genetic variation in a human cohort. Genome Res. 25, 1206–1214 (2015).

77. Maricque, B. B., Chaudhari, H. G. & Cohen, B. A. A massively parallel reporter assay dissects the influence of chromatin structure on cis -regulatory activity. Nat. Biotechnol. 37, 90–95 (2019).

78. Trauernicht, M., Martinez-Ara, M. & Steensel, B. van. Deciphering Gene Regulation Using Massively Parallel Reporter Assays. Trends Biochem. Sci. 45, 90–91 (2020).

79. Kelly Matthew & Semsarian Christopher. Multiple Mutations in Genetic Cardiovascular Disease. Circ. Cardiovasc. Genet. 2, 182–190 (2009).

80. Papaz, T. et al. Factors influencing participation in a population-based biorepository for childhood heart disease. Pediatrics 130, e1198–1205 (2012).

81. Papaz, T. et al. Return of genetic and genomic research findings: experience of a pediatric biorepository. BMC Med. Genomics 12, 173 (2019).

82. Elliott, P. et al. Classification of the cardiomyopathies: a position statement from the European Society Of Cardiology Working Group on Myocardial and Pericardial Diseases. Eur. Heart J. 29, 270–276 (2008).

83. Maron, B. J. et al. Contemporary definitions and classification of the cardiomyopathies: an American Heart Association Scientific Statement from the Council on Clinical Cardiology, Heart Failure and Transplantation Committee; Quality of Care and Outcomes Research and Functional Genomics and Translational Biology Interdisciplinary Working Groups; and Council on Epidemiology and Prevention. Circulation 113, 1807–1816 (2006).

84. Raczy, C. et al. Isaac: ultra-fast whole-genome secondary analysis on Illumina sequencing platforms. Bioinforma. Oxf. Engl. 29, 2041–2043 (2013).

85. Pedersen, B. S. & Quinlan, A. R. Mosdepth: quick coverage calculation for genomes and exomes. Bioinforma. Oxf. Engl. 34, 867–868 (2018).

86. Cingolani, P. et al. A program for annotating and predicting the effects of single nucleotide polymorphisms, SnpEff: SNPs in the genome of Drosophila melanogaster strain w1118; iso-2; iso-3. Fly (Austin) 6, 80–92 (2012).

87. Wang, K., Li, M. & Hakonarson, H. ANNOVAR: functional annotation of genetic variants from high-throughput sequencing data. Nucleic Acids Res. 38, e164 (2010).

88. Danecek, P. et al. The variant call format and VCFtools. Bioinforma. Oxf. Engl. 27, 2156–2158 (2011).

89. Landrum, M. J. et al. ClinVar: public archive of interpretations of clinically relevant variants. Nucleic Acids Res. 44, D862–868 (2016).

90. Stenson, P. D. et al. The Human Gene Mutation Database: towards a comprehensive repository of inherited mutation data for medical research, genetic diagnosis and next-generation sequencing studies. Hum. Genet. 136, 665–677 (2017).

91. 1000 Genomes Project Consortium et al. A global reference for human genetic variation. Nature 526, 68–74 (2015).

92. Fu, W. et al. Analysis of 6,515 exomes reveals the recent origin of most human protein-coding variants. Nature 493, 216–220 (2013).

93. Lek, M. et al. Analysis of protein-coding genetic variation in 60,706 humans. Nature 536, 285–291 (2016).

94. Ng, P. C. & Henikoff, S. SIFT: Predicting amino acid changes that affect protein function. Nucleic Acids Res. 31, 3812–3814 (2003).

95. Adzhubei, I., Jordan, D. M. & Sunyaev, S. R. Predicting functional effect of human missense mutations using PolyPhen-2. Curr. Protoc. Hum. Genet. Chapter 7, Unit7.20 (2013).

96. Schwarz, J. M., Cooper, D. N., Schuelke, M. & Seelow, D. MutationTaster2: mutation prediction for the deep-sequencing age. Nat. Methods 11, 361–362 (2014).

97. Reva, B., Antipin, Y. & Sander, C. Predicting the functional impact of protein mutations: application to cancer genomics. Nucleic Acids Res. 39, e118 (2011).

98. Hubisz, M. J., Pollard, K. S. & Siepel, A. PHAST and RPHAST: phylogenetic analysis with space/time models. Brief. Bioinform. 12, 41–51 (2011).

99. Davydov, E. V. et al. Identifying a high fraction of the human genome to be under selective constraint using GERP++. PLoS Comput. Biol. 6, e1001025 (2010).

100. McLaren, W. et al. Deriving the consequences of genomic variants with the Ensembl API and SNP Effect Predictor. Bioinforma. Oxf. Engl. 26, 2069–2070 (2010).

101. Li, Q. & Wang, K. InterVar: Clinical Interpretation of Genetic Variants by the 2015 ACMG-AMP Guidelines. Am. J. Hum. Genet. 100, 267–280 (2017).

102. Zhu, M. et al. Using ERDS to infer copy-number variants in high-coverage genomes. Am. J. Hum. Genet. 91, 408–421 (2012).

103. Abyzov, A., Urban, A. E., Snyder, M. & Gerstein, M. CNVnator: an approach to discover, genotype, and characterize typical and atypical CNVs from family and population genome sequencing. Genome Res. 21, 974–984 (2011).

104. Poplin, R. et al. Scaling accurate genetic variant discovery to tens of thousands of samples. bioRxiv 201178 (2018) doi:10.1101/201178.

105. Shihab, H. A. et al. Ranking non-synonymous single nucleotide polymorphisms based on disease concepts. Hum. Genomics 8, 11 (2014).

106. Caulfield, M. et al. The National Genomics Research and Healthcare Knowledgebase. (2019) doi:10.6084/m9.figshare.4530893.v5.

107. Raudvere, U. et al. g:Profiler: a web server for functional enrichment analysis and conversions of gene lists (2019 update). Nucleic Acids Res. 47, W191–W198 (2019).

108. Gao, J., Collyer, J., Wang, M., Sun, F. & Xu, F. Genetic Dissection of Hypertrophic Cardiomyopathy with Myocardial RNA-Seq. Int. J. Mol. Sci. 21, (2020).

109. Kim, D., Langmead, B. & Salzberg, S. L. HISAT: a fast spliced aligner with low memory requirements. Nat. Methods 12, 357–360 (2015).

110. Pertea, M. et al. StringTie enables improved reconstruction of a transcriptome from RNA-seq reads. Nat. Biotechnol. 33, 290–295 (2015).

111. Anders, S. & Huber, W. Differential expression analysis for sequence count data. Genome Biol. 11, R106 (2010).

112. Xie, H. et al. Identification of TBX2 and TBX3 variants in patients with conotruncal heart defects by target sequencing. Hum. Genomics 12, 44 (2018).

113. Livak, K. J. & Schmittgen, T. D. Analysis of relative gene expression data using real-time quantitative PCR and the 2(-Delta Delta C(T)) Method. Methods San Diego Calif 25, 402–408 (2001).

114. Hildebrandt, M. R. et al. Precision Health Resource of Control iPSC Lines for Versatile Multilineage Differentiation. Stem Cell Rep. 13, 1126–1141 (2019).

115. Patel, P., Kuzmanov, U. & Mital, S. Avoiding false discovery in biomarker research. BMC Biochem. 17, 17 (2016).

116. Visonà, S. D. et al. Diagnosis of sudden cardiac death due to early myocardial ischemia: An ultrastructural and immunohistochemical study. Eur. J. Histochem. EJH 62, (2018).

117. Ashuach, T. et al. MPRAnalyze: statistical framework for massively parallel reporter assays. Genome Biol. 20, 183 (2019).

