## Supplementary figures and images for "Whole genome sequencing delineates regulatory and novel genic variants in childhood cardiomyopathy"

### Supplementary Figure 1

#### Supplementary Figure 1

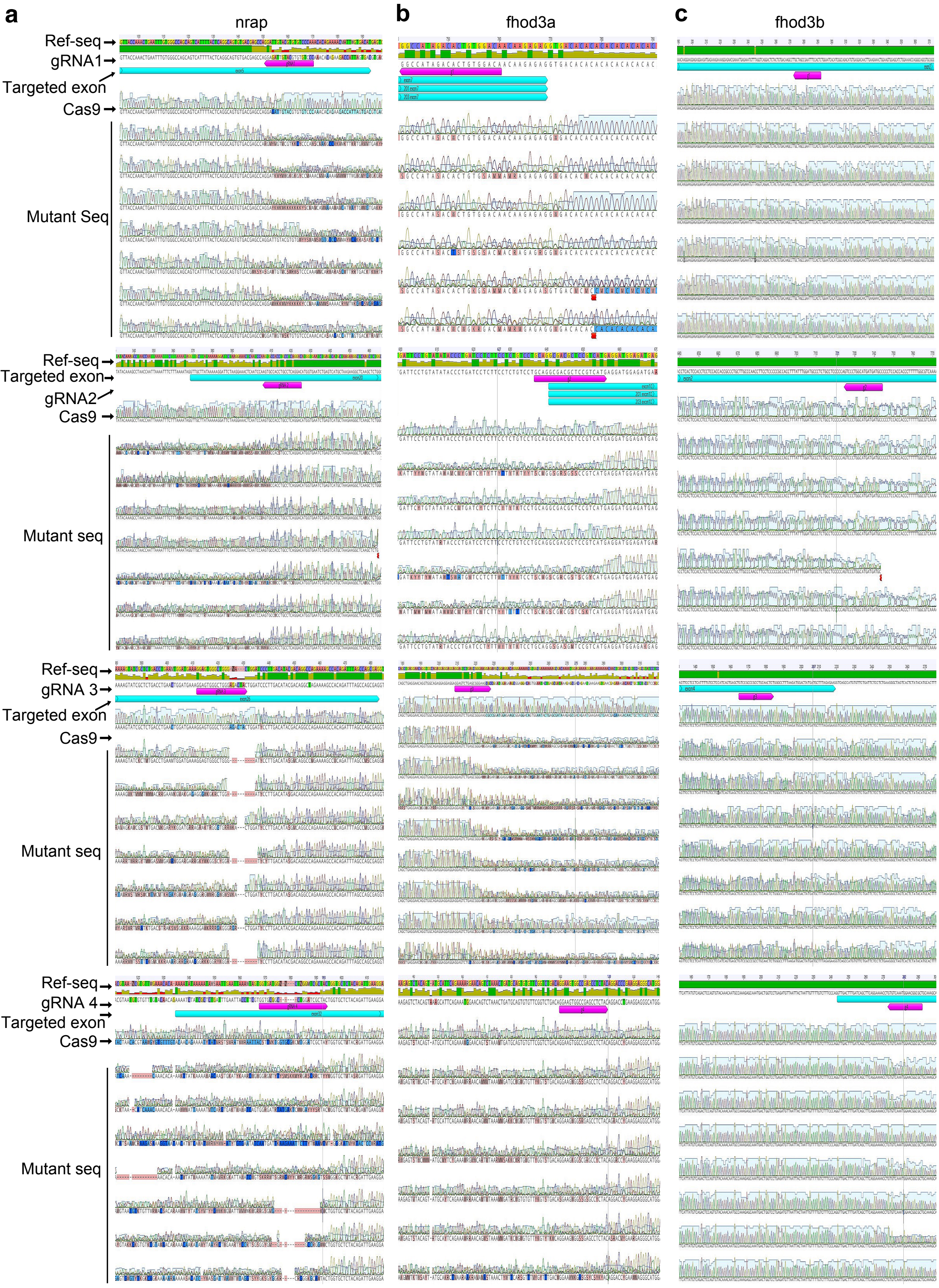

### Supplementary Figure 3

#### Supplementary Figure 3

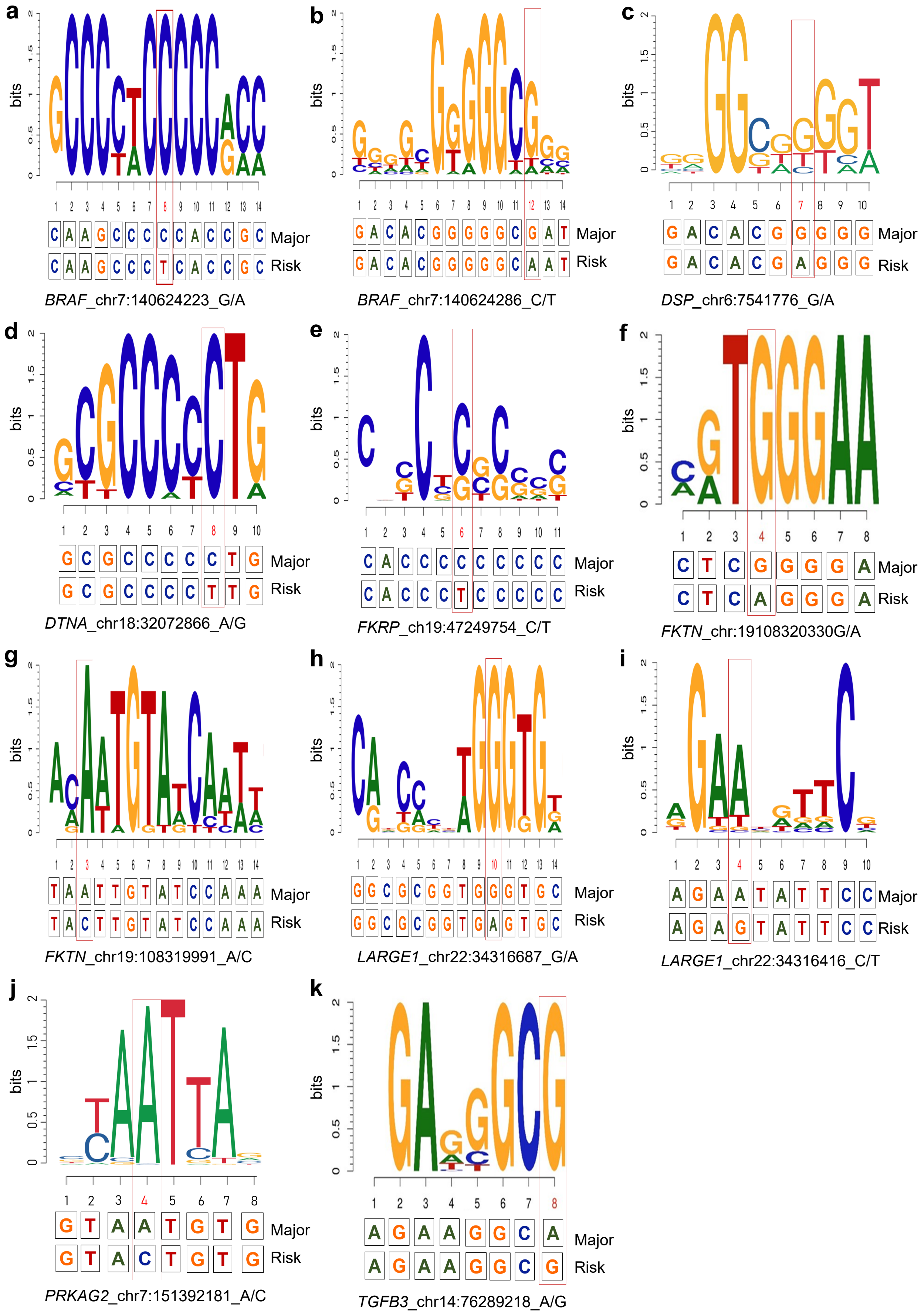

### Supplementary Figure 4

#### Supplementary Figure 4

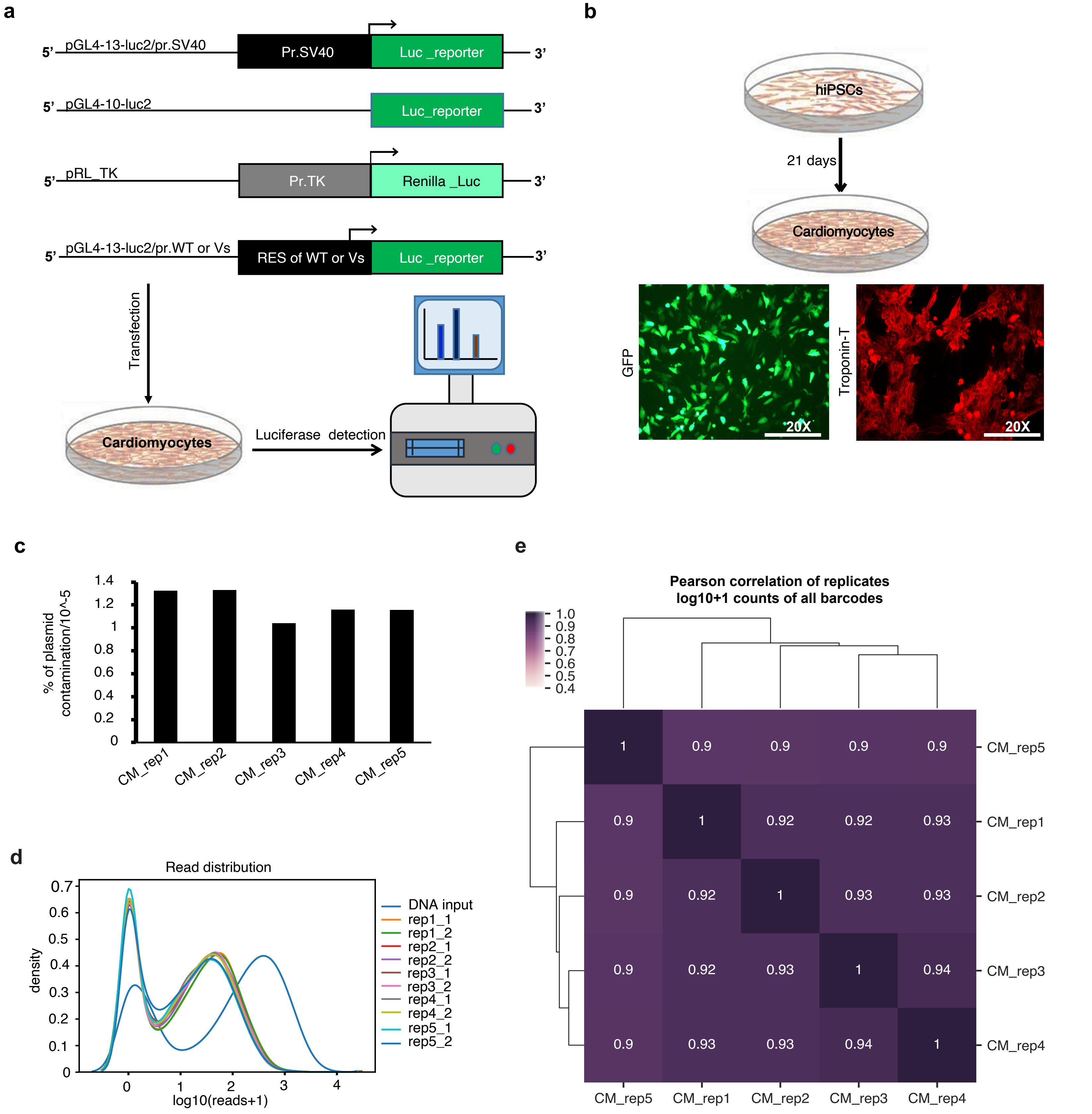
