## Supplementary Figure 2 for "Whole genome sequencing delineates regulatory and novel genic variants in childhood cardiomyopathy"

**a**

### Protein-coding variants

#### Gene Ontology

| Term name | $p_{adj}$ | $-\log_{10}(p_{adj})$ (0 – ≤16) |
|---|---|---|
| actin binding | $1.536\times10^{-10}$ | |
| cytoskeletal protein binding | $2.805\times10^{-10}$ | |
| actin filament binding | $2.630\times10^{-7}$ | |
| structural constituent of muscle | $3.382\times10^{-5}$ | |
| troponin C binding | $5.962\times10^{-5}$ | |
| kinase binding | $7.089\times10^{-4}$ | |
| calmodulin binding | $1.457\times10^{-3}$ | |
| cell adhesive protein binding involved in bundle of His c... | $2.084\times10^{-3}$ | |
| protein-containing complex binding | $3.739\times10^{-3}$ | |
| protein kinase binding | $3.791\times10^{-3}$ | |
| enzyme binding | $1.068\times10^{-2}$ | |
| protein binding involved in heterotypic cell-cell adhesion | $1.619\times10^{-2}$ | |
| ankyrin binding | $2.065\times10^{-2}$ | |

#### Reactome

| Term name | $p_{adj}$ | $-\log_{10}(p_{adj})$ (0 – ≤16) |
|---|---|---|
| Muscle contraction | $6.717\times10^{-11}$ | |
| Striated Muscle Contraction | $5.035\times10^{-10}$ | |
| Cardiac conduction | $3.408\times10^{-2}$ | |
| Ion homeostasis | $4.026\times10^{-2}$ | |

**b**

### Regulatory variants

#### Gene Ontology

| Term name | $p_{adj}$ ↑ | $-\log_{10}(p_{adj})$ (0 – ≤16) |
|---|---|---|
| cell adhesive protein binding involved in bundle of His cell-Purkinje myocyte com... | $1.879\times10^{-9}$ | |
| protein binding involved in heterotypic cell-cell adhesion | $2.672\times10^{-7}$ | |
| cell-cell adhesion mediator activity | $7.553\times10^{-7}$ | |
| cell adhesion mediator activity | $1.745\times10^{-6}$ | |
| cell adhesion molecule binding | $6.107\times10^{-3}$ | |
| FATZ binding | $7.444\times10^{-3}$ | |
| structural constituent of muscle | $1.221\times10^{-2}$ | |
| type III transforming growth factor beta receptor binding | $3.856\times10^{-2}$ | |

#### Reactome

| Term name | $p_{adj}$ ↑ | $-\log_{10}(p_{adj})$ (0 – ≤16) |
|---|---|---|
| Signalling to ERKs | $9.976\times10^{-4}$ | |
| Signaling by FGFR3 | $1.100\times10^{-3}$ | |
| Carnitine metabolism | $1.270\times10^{-3}$ | |
| Signaling by FGFR4 | $1.449\times10^{-3}$ | |
| Signaling by FGFR1 | $2.352\times10^{-3}$ | |
| SOS-mediated signalling | $2.648\times10^{-3}$ | |
| Activated NTRK3 signals through RAS | $2.648\times10^{-3}$ | |
| Activated NTRK2 signals through RAS | $3.969\times10^{-3}$ | |
| SHC-related events triggered by IGF1R | $3.969\times10^{-3}$ | |
| EGFR Transactivation by Gastrin | $5.554\times10^{-3}$ | |
| Signaling by FGFR3 fusions in cancer | $7.401\times10^{-3}$ | |
| MET activates RAS signaling | $7.401\times10^{-3}$ | |
| Signaling by FGFR2 | $9.509\times10^{-3}$ | |
| Activated NTRK2 signals through FRS2 and FRS3 | $9.510\times10^{-3}$ | |
| Constitutive Signaling by Overexpressed ERBB2 | $9.510\times10^{-3}$ | |
| Signaling by FGFR4 in disease | $9.510\times10^{-3}$ | |
| GRB2 events in EGFR signaling | $1.451\times10^{-2}$ | |
| SHC1 events in ERBB4 signaling | $1.451\times10^{-2}$ | |
| SHC-mediated cascade:FGFR3 | $1.451\times10^{-2}$ | |
| Signaling by FGFR | $1.521\times10^{-2}$ | |
| Signaling by NTRK1 (TRKA) | $1.582\times10^{-2}$ | |
| SHC1 events in EGFR signaling | $1.741\times10^{-2}$ | |
| Erythropoietin activates RAS | $1.741\times10^{-2}$ | |
| GRB2 events in ERBB2 signaling | $2.056\times10^{-2}$ | |
| SHC-mediated cascade:FGFR1 | $2.056\times10^{-2}$ | |
| Signaling by EGFRvIII in Cancer | $2.056\times10^{-2}$ | |
| Constitutive Signaling by EGFRvIII | $2.056\times10^{-2}$ | |
| Signaling by NTRK3 (TRKC) | $2.056\times10^{-2}$ | |
| Formation of the cornified envelope | $2.297\times10^{-2}$ | |
| FRS-mediated FGFR3 signaling | $2.397\times10^{-2}$ | |
| SHC-mediated cascade:FGFR4 | $2.764\times10^{-2}$ | |
| Signaling by NTRKs | $3.152\times10^{-2}$ | |
| Gastrin-CREB signalling pathway via PKC and MAPK | $3.157\times10^{-2}$ | |
| SHC-mediated cascade:FGFR2 | $3.157\times10^{-2}$ | |
| FRS-mediated FGFR1 signaling | $3.157\times10^{-2}$ | |
| Tie2 Signaling | $3.157\times10^{-2}$ | |
| Apoptotic cleavage of cell adhesion proteins | $3.157\times10^{-2}$ | |
| Signaling by FGFR3 in disease | $3.576\times10^{-2}$ | |
| Signalling to RAS | $3.576\times10^{-2}$ | |
| Signaling by Ligand-Responsive EGFR Variants in Cancer | $3.576\times10^{-2}$ | |
| Constitutive Signaling by Ligand-Responsive EGFR Cancer Variants | $3.576\times10^{-2}$ | |
| Signaling by FGFR3 point mutants in cancer | $3.576\times10^{-2}$ | |
| Downstream signaling of activated FGFR3 | $4.021\times10^{-2}$ | |
| FRS-mediated FGFR4 signaling | $4.021\times10^{-2}$ | |
| SHC1 events in ERBB2 signaling | $4.021\times10^{-2}$ | |
| FRS-mediated FGFR2 signaling | $4.492\times10^{-2}$ | |
